# A Genetic Model for Central Chondrosarcoma Evolution Correlates with Patient Outcome

**DOI:** 10.1101/2021.11.02.21265785

**Authors:** William Cross, Iben Lyskjær, Tom Lesluyes, Steven Hargreaves, Anna-Christina Strobl, Christopher Davies, Sara Waise, Shadi Hames, Dahmane Oukrif, Hongtao Ye, Fernanda Amary, Roberto Tirabosco, Toby Baker, David Barnes, Christopher Steele, Ludmil Alexandrov, Gareth Bond, Genomics England Research Consortium, Paul Cool, Nischalan Pillay, Peter Van Loo, Adrienne M Flanagan

## Abstract

The treatment options for central chondrosarcoma are limited, and prognoses are generally unreliable. The presence and absence of mutations in *IDH1*, and *IDH2* are defining events, and *TERT* mutations have been recently been associated with poor outcome. Despite this, molecular biomarkers are lacking. Here, analysing data from 356 patients, comprising results from whole genome sequencing (n=68), digital droplet PCR (n=346), and methylation arrays (n=57), we present a comprehensive genetic analysis of chondrosarcoma and suggest its clinical utility. Methylation profiles, *TERT* promoter mutations, genome doubling with prior haploidisation, and age at diagnosis of high grade, distinguish *IDH1-*mutant, *IDH2*-mutant and IDH wildtype tumours. The majority of *IDH2*-mutant tumours harbour *TERT* mutations, though a significant reduction in survival is only found in the less common mutational combination of *IDH1* and *TERT*. We suggest that diagnostic testing for *IDH1, IDH2* and *TERT* mutations could guide clinical monitoring and prognostication.

## Introduction

Central conventional chondrosarcoma (CS) is the most common primary malignant bone tumour in adults. Anatomical location, histopathology, and grading are the current criteria for determining treatment(Bovee et al., 2020; Giuffrida et al., 2009), although these assessments are unreliable for providing prognoses^1–3^, hence a greater understanding of the disease and its biomarkers are required to provide patients with a more personalised treatment plan. Patients with CS grade (G) 1, and cartilaginous tumours of the extremities, irrespective of tumour grade, rarely metastasise or die of their disease. G0/1 tumours that occur in sites where complete excision is difficult, including the axial skeleton and pelvis, are the exception; here tumours often recur and are associated with transformation to higher tumour grade, with many patients eventually succumbing to their disease^4,5^. G2 represents approximately 40% of central CS with a five-year survival of approximately 70%-99%, whereas G3, about 10% of all central CS, have a five year survival of approximately 30-77%^4,5^. The most aggressive form of the disease is the dedifferentiated (DD) subtype, which arises on the background of about 10% of all conventional central chondrosarcomas and has a 5-year survival of 7-24%^2,4,5^. These data highlight the need for improved risk stratification and greater prognostic accuracy.

The cytosolic isocitrate dehydrogenase type 1 (*IDH1*) and mitochondrial isocitrate dehydrogenase type 2 (*IDH2*) enzymes are key components in the tricarboxylic acid cycle. Specific alterations at the R132 and R172 amino acid residues of these genes respectively occur in CS (amongst other cancers), disrupting normal functions and leading to the accumulation of 2-D-hydroxyglutarate (2HG), a competitive inhibitor of alpha KG-dependent dioxygenases. This results in downstream affects, including hypermethylation of CpG islands^6^.

Close to 70% of central CS harbour an *IDH1* (60%) or an *IDH2* (10%) mutation^7^ and these are considered key initiators of disease^7–10^. No recurrent initiating genetic drivers have been reported in the remaining 30% of *IDH1/2* wild type (IDHwt) cases^11,12^, although these tumours have been reported to exhibit different methylation profiles compared to *IDH1* and *IDH2*-mutant tumours^13,14^, hereafter referred to as *IDH1* and *IDH2* tumours. Other key drivers include mutations in *COL2A1, CDKN2A/B, TP53*^11,12,15,16^ as well as less common pathogenic alterations in cell cycle-related genes such as *RB1*^17^ and *CDK4*/*6*^11,18^, alterations in the Indian Hedgehog pathway, and amplifications of *MYC*^11,12^. Alterations in *CDKN2A* and *TP53* are more likely to occur in high grade disease (G2, G3 and DD CS) ^11,16,18^. These alterations have limited value as markers of survival or risk stratification for disease progression. Previously, near haploid karyotypes have been reported in CS^19–21^, although the relationship with other mutations have not been reported.

The recently identified alterations in the *TERT* promoter locus (C228T), which are thought to result in increased telomerase expression leading to immortalisation, rarely occur in well differentiated tumours, suggesting that it likely represents a reliable prognostic marker for CS^22^. In contrast, the impact of *IDH1* and *IDH2* mutations on survival remains unclear^8,23,24^ possibly reflecting the relatively small number of cases studied.

The aim of this study was to undertake a comprehensive analysis of the first large set of chondrosarcoma genomes exploiting data from the Genomics England 100,000 Genomes Project^25^ combined with digital droplet PCR (ddPCR), and methylation profiling. A particular focus of our efforts was to identify mutations of relevance to patient outcome and identify recurrent driver mutations in those tumours that are wild type for *IDH1* and *IDH2* mutations (IDHwt).

## Results

### Driver mutations in central conventional and dedifferentiated chondrosarcomas

Profiling a total of 350 CS cases, including ddPCR (n = 282) and WGS (n = 68), we verified previous findings that *IDH2* are less frequent than *IDH1* mutations (*IDH1*: 51%, *IDH2*: 14%, IDHwt: 35%) ^15,24,26^. In cases with grading information (n = 343) we found that *IDH1* mutations were equally frequent across grades (G0/I: 51%, G2/3: 53%, DD CS: 55%; p = 0.9), *IDH2* mutations were more frequent in higher grade disease (G0/I: 7%, G2/3: 14%, DD: 25%; p = 0.005). IDHwt was negatively associated with increasing grade (%IDHwt; G0/I: 41%, G2/3: 32%, DD CS: 19%; p = 0.01). These data imply that the progression to DD CS is more common in tumours with *IDH2* mutations, and least common in IDHwt tumours.

Canonical mutations and structural changes near the *TERT* promoter have been reported in approximately 20% of CS and found to correlate with high grade disease^22,24,27^. We found a similar frequency in our cohort (23%, 74 with C228T, one with C250T, four with structural changes near the *TERT* promoter (**Supplementary Figure 1**). *TERT* mutations were rare in well differentiated tumours and increased in frequency across grades (G0/I: 3%, G2/3: 22%, DD: 56%; p = 2e-14, **Figure 1A**). We found that the *TERT* promoter was hypermethylated in 19/57 (33%) of cases analysed on methylation arrays, excluding DD CS cases (**Figure 1B**). Seven of these cases, all high grade, also harboured *TERT* C228T promoter mutations. There was no significant difference between the number of cases with both *TERT* promoter mutations and hypermethylation across *IDH1* and *IDH2* tumours (**Figure 1B**). Rare alterations involving *ATRX* have been reported previously^24^ but in the 100KGP cohort (n = 68), though we found no such alterations in this cohort. These data confirm that *TERT* mutations, and possibly methylation, have distinct roles in CS progression.

**Figure 1:**
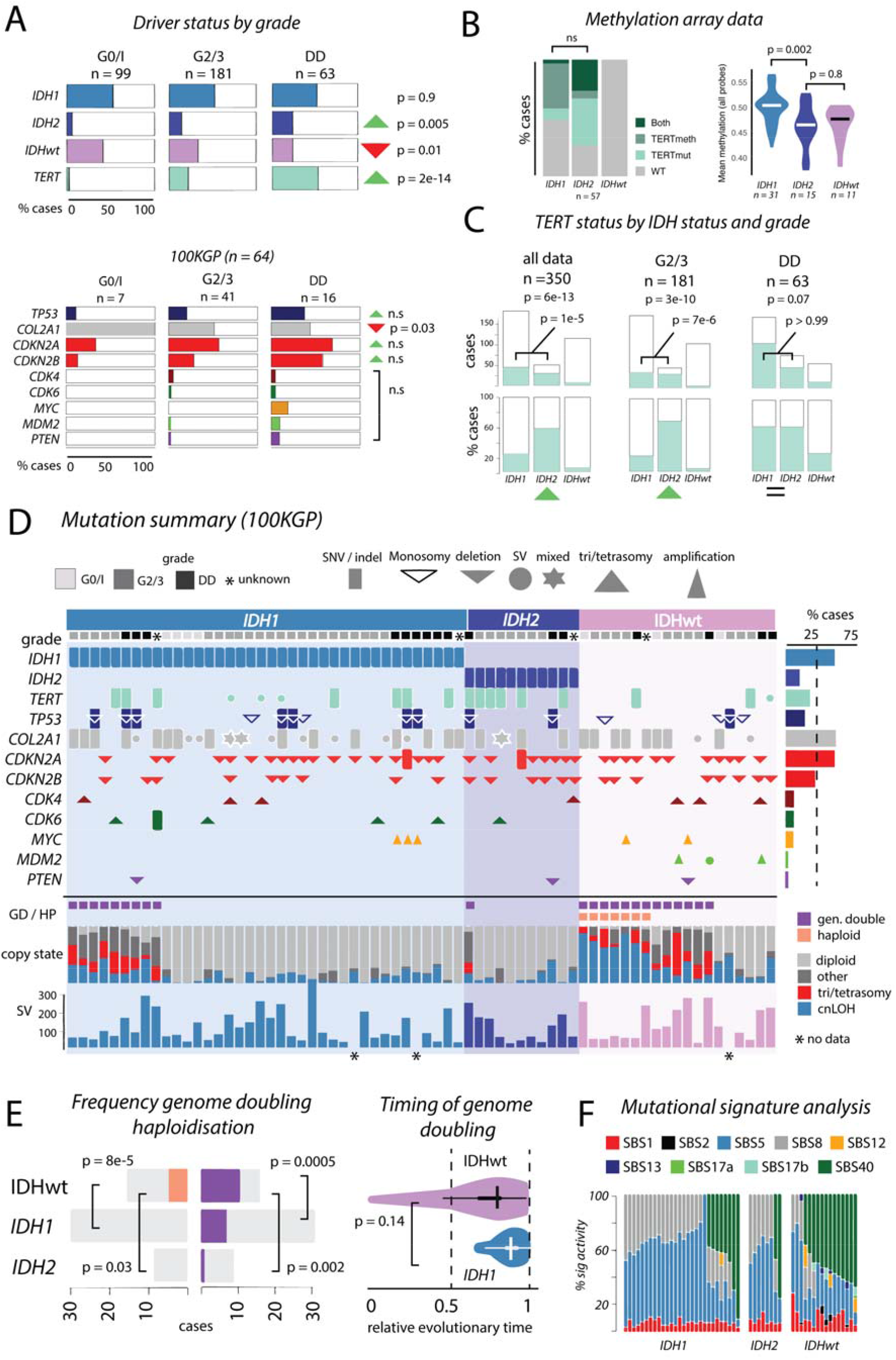
Summary of Genomics England Cohort. **Box A**: Summary of driver mutations by grade. *IDH1* mutations are frequent across all grades, though *IDH2* and *TERT* mutations are enriched in G2/3 and DD CS tumours. **Box B**: *TERT* mutation and methylation status (left) and overall genomic methylation levels (right) across IDH groups. **Box C**: *TERT* mutation status across IDH groups. Canonical *TERT* promoter mutations are common in *IDH2* mutant tumours but rare in IDHwt tumours (left plot). In G2/3 tumours *pTERT* is more common in *IDH2* compared to *IDH1* tumours (middle plot), though equally common in DD CS (right plot). p-values for tests across all IDH groups above, *IDH1* vs *IDH2* are marked on plots. **Box D**: Mutational calling showing driver calls, genome doubling (GD) and haploidisation (HP), Battenberg copy states (diploid, gain, copy neutral LOH, and any other copy state), and Delly structural variant calls (methods). **Box E**: GD and HP overview by IDH status. **Box F**: Mutational signature analysis demonstrating commonality of SBS2, 5, and 8, and prominence of SBS40 in IDH groups.

We next examined mutations in other key driver genes reported in CS, utilising the 100KGP cohort (**Figure 1A)**. Our findings were largely similar to those previously reported^12,28^. Monosomies of 17p and pathogenic SNVs/indels in *TP53* were found in 22% of cases in line with previous reports^11^. *TP53* mutations were absent from all but one well differentiated tumour. *COL2A1* mutations were common, and marginally anti-correlated with increasing grade (G0/I: 100%, G2/3: 54%, DD CS: 44%; p = 0.03). As previously reported^16,18,19^, pathogenic SNVs/indels and/or bi-allelic deletions of *CDKN2A* and *CDKN2B* were common in CS and enriched in G2/3 and DD CS, though not significantly in this dataset. Hypermethylation of these genes was not detected. *CDK4* and *CDK6* gains were found in 12 cases, and a single case had a pathogenic SNV in *CDK6*. These frequencies are similar to previous reports^11^. *MYC* amplifications were found in five high-grade tumours establishing its status as a driver of CS^29,30^. *MDM2* alterations were identified in three high grade IDHwt cases, two of which were amplifications (one 8 copies, one 31 copies, confirmed using fluorescence in situ hybridisation), and one was a structural alteration involving intron 7 of *MDM2* and an intragenic region on chr4q28.3. The latter did not result in amplification of *MDM2* but resulted in removal of the zinc finger binding domains, which has been suggested to have an oncogenic effect^31^. All three mutations were mutually exclusive of *TP53* mutations. These data suggest that *MDM2* mutations can act as drivers in IDHwt CS ^24^. Homozygous deletions of *PTEN* were present in three high grade cases. *PTEN* promoter hypermethylation was found in 13/57 cases, all high grade. Analysis utilising dNdS^32^ returned no previously unknown drivers, implying that all prominent somatically mutated genes driving CS have likely been identified (**Online Methods, Supplementary Figure 2**).

**Figure 2:**
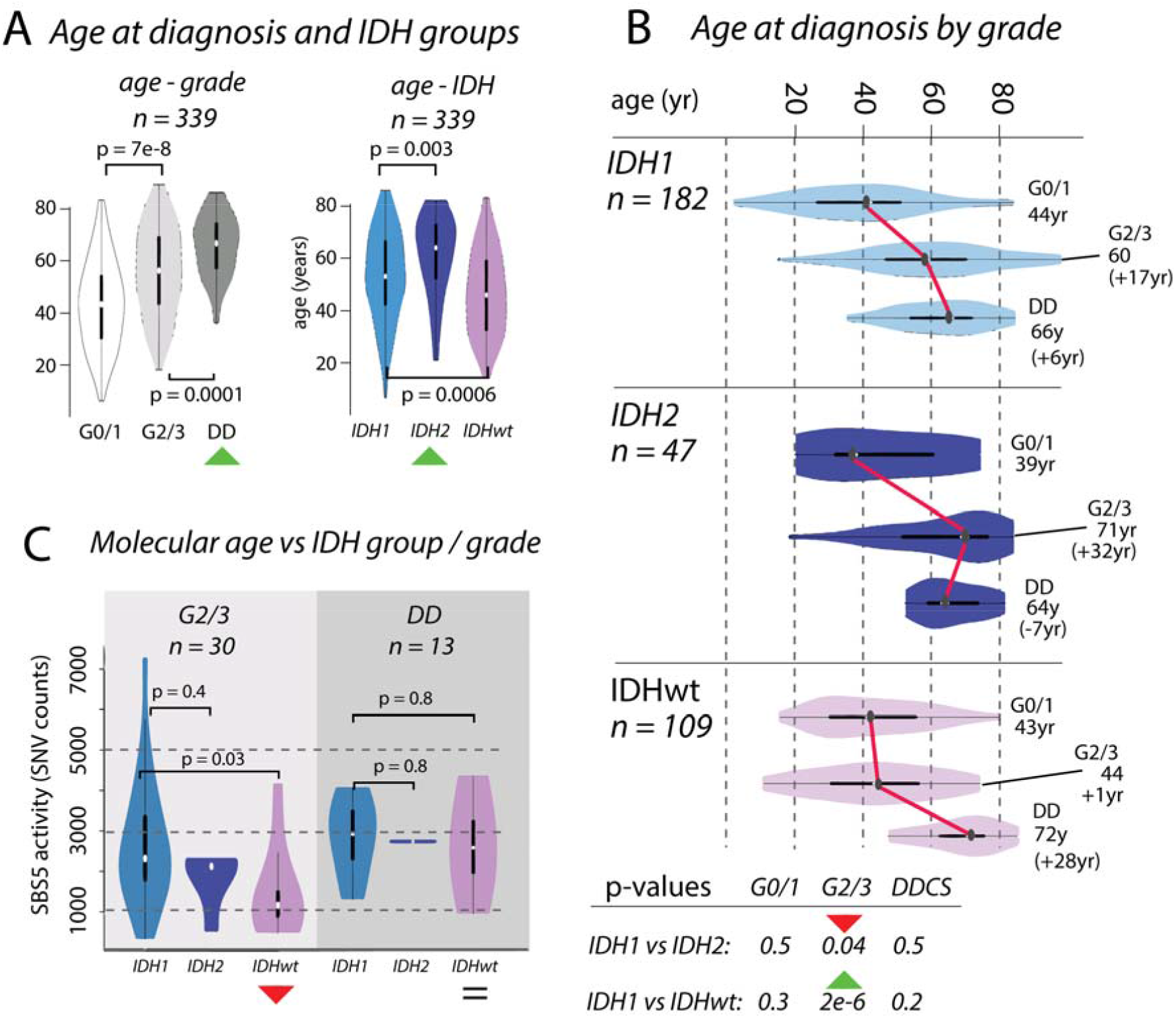
Divergences in chronological and molecular age in chondrosarcoma. **Box A:** Boxplots showing the distribution of age at diagnosis (n = 339) increasing across grades. Distributions differ across *IDH1, IDH2* and IDHwt groups, with *IDH2* tumours occurring in older patients compared to those with *IDH1* and IDHwt tumours. **Box B:** Boxplots of age, broken down by IDH status and grade. **Box C:** The differences in chronological age between G2/3 *IDH1* and *IDH2* tumours and IDHwt tumours (**Box B**) is recapitulated in the activities of mutational signature SBS5. There is no significant difference in molecular age of *IDH1* and *IDH2* tumours, whereas there is a significant difference in the chronological age.

### IDH1, IDH2, and TERT define key genetic subgroups

Analysis of all mutation calls (n = 350) revealed that the frequency of *TERT* mutations was different across *IDH1, IDH2* and IDHwt cases (**Figure 1C**). *IDH2* mutations were strongly associated with *TERT* mutations (IDHwt: 5%, *IDH1*: 24%, *IDH2*: 58%, p = 6e-13; *IDH1 vs IDH2:* p = 1e-5). This association was observed in G2/3 (*IDH1 vs IDH2:* p = 7e-6) but not in DD CS (*IDH1* vs *IDH2*: p > 0.99), implying that although *TERT* is associated with high-grade tumours, this is not equal in the context of IDH mutation status.

### Hypermethylation across IDH1- and IDH2-mutated tumours

CpG island DNA hypermethylation has been reported to distinguish between cartilaginous *IDH* and IDHwt tumours^13,14^. However, utilising the larger numbers available in this study, we found 3,468 differentially methylated probes (DMPs) across *IDH1* and *IDH2* tumours, excluding DD CS (n = 31, p = 0.002, **Supplementary Figure 3**). The overall methylation level across all probes also revealed significant differences between *IDH1* and *IDH2* tumours (p = 0.002) indicating that the former are globally hypermethylated compared to *IDH2* and IDHwt tumours.

**Figure 3:**
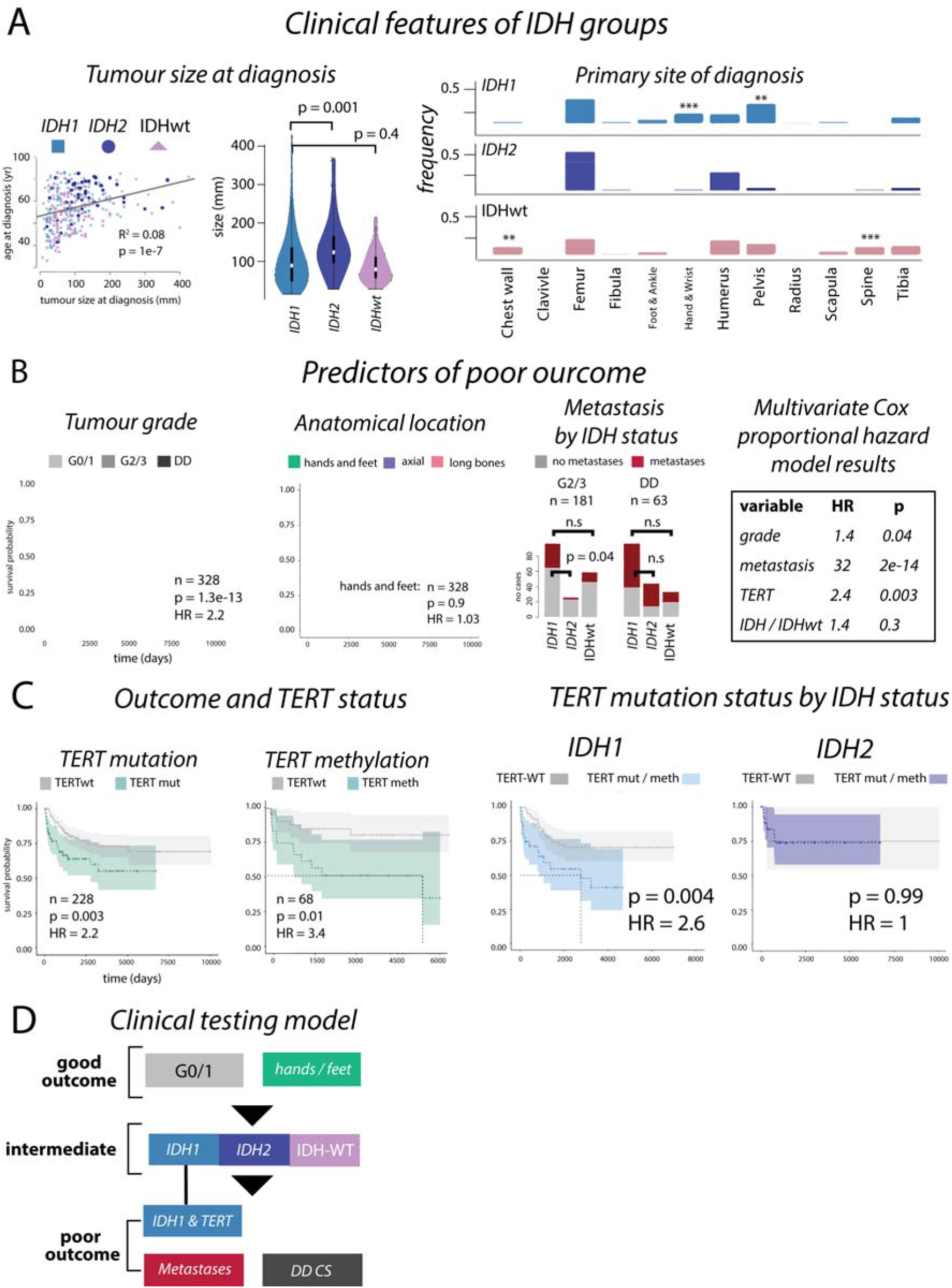
Outcomes in chondrosarcoma. **Box A:** Size of *IDH1, IDH2* and IDHwt CS are different at presentation and their anatomical location are largely comparable, although IDHwt tumours develop more frequently in the chest wall and spine (left and middle, and right, respectively). **Box B:** Kaplan-Meier analysis and hazard ratios (HR) from Cox proportional hazard analysis confirms tumour grade, and anatomical location, as predictors of outcome. The frequency of metastasis is significantly lower in *IDH2* G2/3 disease compared to *IDH1* G2/3 disease but is comparable in DD CS. **Box C:** *TERT* mutations are linked to poor outcome, as is methylation of *TERT* (left plots). In high-grade (G2/3 and DD CS) *IDH1* tumours, *TERT* mutations associate with a poor outcome, but not in *IDH2* tumours. **Box D:** A proposed clinical testing model to determine outcome in central chondrosarcoma.

### Partial haploidisation followed by genome doubling is common in IDHwt tumours

We compared the mutational profiles across each IDH group (complete summary of 100KGP data shown in **Figure 1D**) and found that the frequency of common drivers, excluding *TERT*, was similar across *IDH1* and *IDH2* and IDHwt tumours. We did not find that mutations in *CDKN2A/B and TP53* were enriched in *IDH1/2* cases, as previously reported^33^ but contrasting another study^24^. The total number of SVs was not statistically different across the *IDH* groups, nor was the number of SVs that fell into gene regions. We found 32 genes harboured SVs in approximately 25% of cases, although none of these were cancer-related genes (**Online Methods**).

The genetic alterations initiating development of IDHwt CS remains unknown, but previous reports of near-haploid (HP) and hyperhaploidy in chondrosarcoma and in other sarcoma subtypes including undifferentiated sarcomas and malignant peripheral nerve sheath tumours, prompted us to investigate^19,20,34,35^. We found 23 tumours with genome doubling (GD) and seven with HP in the 100KGP cohort (n=68, **Supplementary Figure 4, Supplementary Methods**). Most GD events (16/23, 69%) occurred in the absence of HP, whereas HP always occurred with GD (**Figure 1D)**. GD was highly enriched in IDHwt tumours (GD%, *IDH1*: 24%, *IDH2*: 9%, IDHwt: 63%, IDHwt vs *IDH1* p = 0.0005, **Figure 1E**), and HP was exclusive to this group (HP%, *IDH1*: 0%, *IDH2*: 0%, IDHwt: 37%, IDHwt vs *IDH1* p = 8e-5, **Figure 1E**). Timing analysis demonstrated that GD events tended to occur at a similar relative time in IDHwt and *IDH1* cases implying that it could be an intermediate or late event in evolutionary timelines of both tumour groups (**Figure 1E, Online Methods)**. The six cases of IDHwt tumours without HP/GD events, harboured mutations in *TP53* and *CDKN2A*, although alterations in these genes were not mutually exclusive with the absence of GD and HP (*TP53*: 3/6, 50%, *CDKN2A/B*: 5/6, 83%, **Figure 1D**). One of these cases was G0/1, pointing to a possible initiating role of *TP53* and *CDKN2A* in some IDHwt tumours.

### Mutational signatures across IDH1, IDH2, and IDHwt groups

Analysis of mutational signatures in the 100KGP cohort (n = 52, **Figure 1F, Supplementary Figure 5**) revealed nine active signals, with SBS1, SBS5, and SBS8 being ubiquitous and most prominent across *IDH1, IDH2*, and IDHwt tumours. Five signatures (SBS2, SBS12, SBS13, and SBS17a/b) were principally exclusive to IDHwt tumours. SBS2 and SBS13 have been associated with APOBEC and were simultaneously active in five IDHwt cases (18%). We did not observe any difference in SNV burden in tumours with active SBS2 and SBS13. SBS12, was found in one *IDH1* case and three IDHwt cases. SBS17a/b, signatures with unknown aetiology, were found only in IDHwt cases. SBS40, also of unknown aetiology, was found in 28% of *IDH1* cases, 25% of *IDH2* cases, and 81% of IDHwt. These data demonstrate that *IDH1* and *IDH2* tumours are comparable in terms of mutational signatures, whereas IDHwt tumours exhibit more heterogeneous mutational processes.

### The genetic distinction between central conventional and dedifferentiated chondrosarcoma

We next analysed the DD CS for specific alterations that may explain their histological phenotype and their poor clinical outcomes. We confirmed that metastatic disease was most common in DD CS (60%, compared to 27% in G2/3 and <1% in G0/1 (G2/3 vs DD: p = 1e-5). Analysing the 100KGP data (DD: n = 16, G2/3: n = 41), the frequency of identified known drivers in DD CS and G2/3 revealed no difference except for *IDH2* and *TERT*, which were enriched in DD (*IDH2*: p = 0.05, *TERT*: p = 3e-6). However, we found differences in total driver burden (p = 2e-8), SNV burden (p = 0.009), number of chromosome segments (p = 0.01) and SV burden (p = 0.01) (**Supplementary Figure 6**). We next explored whether the increased segment counts were attributable to chromothripsis. Using a previously published method^36^ we found only one instance of chromothripsis (WGS_21) which overlapped with the SV identified at the *TERT* loci (Figure 1D, Supplementary Figure 1). Examining the number of more broadly, the average number of chromosomes with high breakage was higher in DD CS compared to G2/3 (median, G2/3: 0, DD CS: 2.5, p = 0.03, see **Online Methods**). There were no specific chromosome arms enriched among those with high fragmentation, although three cases (19%) had fragmentation across chromosome 12q, which has also been reported in dedifferentiated liposarcoma^37^. Previous studies have reported that aberrations of chromosome 5q and trisomy of chromosome 19 distinguish G2/3 from DD CS^38^. 25% DD CS harboured 19p/q gains which is less than the 50% previously reported^38^. Examining losses and gains across all chromosome arms revealed no events unique to DD CS although losses at 15q were more common in this subtype (15q loss, G2/3: 10%, DD CS: 38%, p = 0.05). Together these analyses suggest that the primary genetic difference between G2/3 and DD CS is the number of accrued SNVs and the degree chromosome fragmentation.

### Age at diagnosis as a clinical factor in chondrosarcoma

Previous studies of CS have treated *IDH1* and *IDH2* tumours as one group ^13,39^. Our results, leveraging hundreds of cases, provide evidence that *IDH1* and *IDH2* mutations lead to distinct genetic pathways, with differences in the frequency of *TERT* mutations, GD/HP, methylation profiles, and the number and types of mutational signatures.

We examined the effect of *IDH1* and *IDH2* mutations and the absence of these mutations on the clinical behaviour of CS (n = 339, **Figure 2**). We showed that patient age at diagnosis increased across grades in these groups, and that the median age was highest in those with *IDH2* tumours (*IDH1*: 55yr, *IDH2*: 67yr, IDHwt: 47yr, *IDH1* vs *IDH2*: p = 0.003, *IDH1* vs IDHwt: p = 0.0006, **Figure 2A**). The age at diagnosis for each IDH group was similar for G0/1 and DD CS, and the difference in age of the G2/3 tumours explained the overall difference in ages (median age G2/3, *IDH1*: 60yr, *IDH2*: 71yr, IDHwt: 44yr, *IDH1*vs *IDH2*: p = 0.04, *IDH1* vs IDHwt: p = 2e-6, **Figure 2B**). We considered whether these differences in chronological age at diagnosis were reflected in the mutational signatures active in each group. The total SNV burden correlated with age at diagnosis, as did SBS5, previously been reported as clock-like^40^, SBS8, but not SBS1. In G2/3 tumours, the activity of SBS5 was similar in *IDH1* and *IDH2*, but lower in IDHwt (*IDH1* vs *IDH2* p = 0.4, *IDH1* vs IDHwt: p = 0.03, **Figure 2C**). By contrast, SBS5 activity was similar in all DD cases (*IDH1* vs *IDH2* p = 0.7, *IDH1* vs IDHwt: p = 0.8, **Figure 2C**). These same results were recapitulated when using SBS8 and total SNV burden (**Supplementary Figure 7**). Together, these data imply further differences in the rate of evolution from G2/3 to DD CS across *IDH1, IDH2*, and IDHwt tumours.

### Divergent outcomes in IDH1, IDH2 and IDHwt tumours

Using all available clinical information (n = 342) we found that *IDH2* tumours tended to be larger at time of presentation (*IDH1* vs *IDH2*: p = 0.001, *IDH1* vs IDHwt: p = 0.4, **Figure 3A**), supporting the premise that these tumours evolve over longer time periods, and present in older people. Development in specific anatomical locations was not significantly different (**Figure 3A**).

Using all cases with available follow-up data (n = 328), a Cox proportional hazard model demonstrated that G0/1 tumours nearly always had a good outcome with no metastatic events being recorded and only one of 98 patients, with a pelvic tumour, succumbing to disease. As expected, no patients with tumours in the small bones of the hands and feet died of disease (**Figure 3B**). We found that DD CS had a higher frequency of metastatic disease compared with G2/3 disease (G2/3 vs DD CS, p = 9e-7). There were no significant differences in the frequency of metastatic disease across *IDH1, IDH2* and IDHwt DD CS tumours. However, metastases appeared to occur less frequently in patients with *IDH2* G2/3 tumours compared to *IDH1* and IDHwt tumours (% metastases, *IDH1*: 37%, *IDH2*: 13%, IDHwt: 23%, p = 0.06, *IDH1* vs *IDH2*: p = 0.04, **Figure 3B**). As expected, survival of all high-grade disease (excluding small bones of the hands and feet) were significantly influenced by metastasis (univariate HR = 30, p = 4e-13). Multivariate Cox proportional hazard analysis of metastasis, grade, *TERT* mutation status, and *IDH* mutations in tumours of the long bones and axial skeleton demonstrated that all factors, except IDH status, significantly affected outcome (**Figure 3B**).

Canonical *TERT* promoter mutations (g.1295113) had an independent hazard ratio (HR) that was equal to that of grade (*TERT*: HR = 2.2, p = 0.003, tumour grade: HR = 2.2, p = 2e-13, **Figure 3B, C, Supplementary Figure 8**), pointing to the benefit of *TERT* as a biomarker. We also found that overall outcomes were worse in patients whose tumours had *TERT* hypermethylation (n = 68, HR = 3.4, p = 0.01). Restricting our analyses to high-grade tumours and excluding tumours in the hands and feet, we found that patients whose tumour harboured both *IDH1* and *TERT* mutations had significantly worse outcomes than those with an *IDH1* mutation alone. *TERT* mutations had no effect on outcome in patients with *IDH2* tumours, even though these mutations are found more frequently in combination with *IDH2* mutations (**Figure 3C, Online Methods**). This suggests that *TERT* mutations are context specific and only relevant to outcome predictions in *IDH1* tumours. Given this finding, and the context of our other results, we suggest a clinical risk assessment model for *IDH* and *TERT* mutation status that could aid prognostication (**Figure 3D**).

## Discussion

In this study of patients with CS, involving targeted, whole genome, and methylation data, we have obtained significant insights into the genetic pathways and dynamics underlying disease progression. Here, with the benefit of greater sample numbers, we have been able to study tumours with *IDH1* and *IDH2* mutations independently and shown that they represent distinct genetic and clinical groups. In addition to confirming that *IDH2* tumours represent the minority group with 14% of cases, we report that they present as larger tumours and on average over a decade later than *IDH1* tumours. Despite this, *IDH1* and *IDH2* tumours have similar molecular ages which suggests that on average, tumours with *IDH2* mutations have slower cell division rates. Therefore, we speculate that many of these tumours go into growth arrest and become calcified, representing at least a proportion of calcified enchondromas, a lesion commonly detected, when medical imaging is undertaken for unrelated symptoms. This would also account for the comparatively lower frequency of these tumours. Furthermore, the high incidence of p*TERT* mutations and or high levels of promoter methylation in high grade *IDH2* tumours, suggests that these events, through activation of telomerase, have prevented the senescent phenotype and bring about high-grade *IDH2* tumours. This finding could also account for the presentation at the relatively late age of these tumours.

Apart from the presence of *IDH1* and *IDH2* mutations, no significant differences in the type or number of mutations were identified that accounted for the different clinical findings associated with these tumour groups. However, we show here that *IDH1* tumours are globally more methylated at CpG islands compared to both the *IDH2* and IDHwt tumours. The small numbers of cases studied to date is likely to account for this being unrecognised previously^13,14^. Indeed, even *IDH1* and *IDH2* gliomas, which are considerably more common than CS, are generally studied together because of their small numbers. Nevertheless, although all *IDH* mutations result in accumulation of 2-HG there is growing evidence that the impact of the different mutations exerts different biological effects. Studies utilising human oligodendroglioma cells have shown that the *IDH1* R132 mutation leads to higher enzymatic activity than that brought about by *IDH2* R172^41^. Other studies of *IDH1* and *IDH2* mutations in gliomas point to them as having distinct mutational and clinical patterns^42^. Furthermore, different biological effects of 2HG are also seen as a consequence of different *IDH2* mutations(Kotredes et al 2019). As it is known that 2HG exerts diverse biological functions including regulation of DNA hydroxymethylation, it is feasible that the *IDH1* and *IDH2* mutations explain our different methylation array findings and mediate the different behaviour of the tumour subgroups. However, further research is required to establish this.

Previous studies have suggested different effects of *IDH* mutations on clinical outcome in patients with CS^8,23,24^. Here, with the benefit of a large patient cohort, we show that although *IDH2* tumours are more commonly associated with *TERT* mutations, only *IDH1* mutations in combination with *TERT* mutations are associated with significantly reduced survival. We also highlight that identification of structural alterations and hypermethylation of this gene is useful for increasing prognostic accuracy.

Based on our findings, we propose a risk stratification model for patients with central CS. Our recent study showing that detection of mutant IDH molecules in the circulation correlates with a poor prognosis could be used in conjunction with the proposed model^43^.

## Methods

### Patients and samples

356 cases of CS were included in the study. These included 68 tumour-normal paired samples from four clinical sites that were subjected to whole genome sequencing as part of the Genomics England 100,000 Genomes Project (hereby referred to as the *100KGP* cohort, **Supplementary Table 1, Supplementary Table 2**). The remaining cases were obtained from the archives of the Royal National Orthopaedic Hospital (RNOH, Stanmore, UK)(**Supplementary Table 3**). We analysed 84 cases using methylation arrays (**Supplementary Table 4, Online Methods**).

### Bioinformatic pre-processing and statistical assessment

Single nucleotide variants (SNVs) and indels were called on whole-genome sequencing (WGS) data via Strelka and filtered using a panel of normal (PON) samples ^44^ (**Online Methods**). To quality assess the 100KGP mutation calls, we performed orthogonal verification of hotspot mutations in *IDH1* (R132), *IDH2* (R172) and *TERT* (C228T) identified across 64 patients (59 mutations in total) using ddPCR, which yielded a recall rate of 100%. There was one instance (WGS_53) where a mutation was called by ddPCR but not in the WGS data (*IDH1* ddPCR, IDHwt WGS, later result used, **Online Methods**). Somatic copy number variants were called using Battenberg^45^ and structural variants (SVs) were called using Delly (v0.8.5) ^46^. Unless otherwise specified comparisons between groups were performed using Wilcoxon tests for distributions and Fisher exact tests for group counts (i.e. in *IDH1*/*2*/WT group comparisons). Survival analysis utilised a Kaplan-Meier standard Cox proportional hazard model.

### Identification of driver mutations, genome doubling, partial haploidisation, and analysis of mutational signatures

Driver mutations in SNVs and indels were identified using a combination of known hotspot locations published previously and available online (**Online Methods**), the SIFT^47^ and POLYPHEN^48^ tools, plus visual inspection using integrative genomics viewer (IGV) of the *IDH1* R132, *IDH2* R172 and *TERT* mutations (**Supplementary Note 1**). Amplification events were designated as copy states of five or in diploid genomes, and nine or more in those that are genome doubled. For the purposes of plotting, we classified copy states from Battenberg as either, diploid, trisomy or tetrasomy, copy neutral LOH (cnLOH), and the remaining copy states as “other” (any remaining copy state). Tumours with genome doubling were identified using a clustering procedure based on the R package Mclust^49^. Cases with more than 50% LOH were marked as exhibiting partial haploidisation. We confirmed the ploidy status in 14 of the 100KGP cases using flow cytometry (**Online Methods, Supplementary Note 2**). To time the appearance of genome doubling we used a methodology based on molecular-clock principles^50,51^. 96 channel single-base substitution mutational signatures were extracted from the Strelka-called SNVs using SigProfilerExtractor^52^ version 1.1.3 with default parameters.

### Methylation data protocol and analysis

500 ng of DNA from frozen tumour samples were bisulphite converted using Zymo EZ DNA methylation Gold kit (Zymo Research Corporation Irvine, CA, USA) and hybridised to the Infinium HumanMethylationEPIC beadchip arrays (Illumina, San Diego, CA). The generated methylation data were analysed using the ChAMP R^53^, normalised using BMIQ and hierarchical clustering plots were constructed using the ‘pheatmap’ R package^54^.

*TERT* promoter methylation status was determined by the methylation status of the cg11625005 probe as reported previously^55^. Raw DNA methylation data files have been deposited in the ArrayExpress database (www.ebi.ac.uk/arrayexpress, accession: E-MTAB-11031).

### Statistics and mathematical analysis

In all group comparison situations, such as *IDH1, IDH2*, IDH-WT cases, with or without *TERT* mutations, we used Fisher test statistics as implemented in R (testing both 3×2 and 2×2 contingency tables). Distributions of data, as seen in the tests of timing for genome doubling (Figure 1E) was performed using Wilcoxon tests, again implemented in R. Linear regressions (Figure 3A) were also implemented using the standard R methods. The cox proportional hazard model and Kaplan-Meyer analyses were performing using the *survminer* package^56^. For the chromosome arm frequency comparisons, we used fisher tests and the Bonferroni multiple testing correction. For power calculations please see online methods.

## Supporting information

Supplementary figures and stats

full methods

## Data Availability

All data produced are available via the supplementary material or online following application to Genomics England.

## Data availability

All ddPCR results are included in Supplementary table 3. The SNV, indel and structural calls produced from the WGS data are available in Supplementary table 2. The bam and vcf files of these data are available on application for access to Genomics England and the Clinical Interpretation Partnership. The raw methylation array files are available via the ArrayExpress database (www.ebi.ac.uk/arrayexpress, accession: E-MTAB-11031)

## Acknowledgements

Funding for this study was received from Sarcoma UK (SUKG01.18), Bone Cancer Research Trust infrastructure grants to AMF and PC, RNOH NHS R&D grant (AMF). IL is supported by the Lundbeck Foundation (grant: R303-2018-3018). The project was also supported by the National Institute for Health Research, UCLH Biomedical Research Centre, the UCL Experimental Cancer Centre, and the Francis Crick Institute, which receives its core funding from Cancer Research UK (FC001202), the UK Medical Research Council (FC001202), and the Wellcome Trust (FC001202). For the purpose of open access, the authors have applied a CC BY public copyright licence to any author accepted manuscript version arising from this submission. PVL is a Winton Group Leader in recognition of the Winton Charitable Foundation’s support towards the establishment of The Francis Crick Institute. We are grateful to the Biobank Team at the RNOH and RJAH, and all healthcare workers who cared for the patients. We specifically wish to thank the patients for engaging in our research.

We would like to acknowledge the work of the Genomics England research consortium: Ambrose, J. C.^1^; Arumugam, P.^1^; Bevers, R.^1^; Bleda, M.^1^; Boardman-Pretty, F.^1,2^; Boustred, C. R.^1^; Brittain, H.^1^; Caulfield, M. J.^1,2^; Chan, G. C. ^1^; Fowler, T. ^1^; Giess A.^1^; Hamblin, A.^1^; Henderson, S.^1,2^; Hubbard, T. J. P.^1^; Jackson, R.^1^; Jones, L. J.^1,2^; Kasperaviciute, D.^1,2^; Kayikci, M.^1^; Kousathanas, A.^1^; Lahnstein, L.^1^; Leigh, S. E. A.^1^; Leong, I. U. S.^1^; Lopez, F. J.^1^; Maleady-Crowe, F.^1^; McEntagart, M.^1^; Minneci F.^1^; Moutsianas, L.^1,2^; Mueller, M.^1,2^; Murugaesu, N.^1^; Need, A. C.^1,2^; O‘Donovan P.^1^; Odhams, C. A.^1^; Patch, C.^1,2^; Perez-Gil, D.^1^; Pereira, M. B.^1^; Pullinger, J.^1^; Rahim, T.^1^; Rendon, A.^1^; Rogers, T.^1^; Savage, K.^1^; Sawant, K.^1^; Scott, R. H.^1^; Siddiq, A.^1^; Sieghart, A.^1^; Smith, S. C.^1^; Sosinsky, A.^1,2^; Stuckey, A.^1^; Tanguy M.^1^; Taylor Tavares, A. L.^1^; Thomas, E. R. A.^1,2^; Thompson, S. R.^1^; Tucci, A.^1,2^; Welland, M. J.^1^; Williams, E.^1^; Witkowska, K.^1,2^; Wood, S. M.^1,2^

1. Genomics England, London, UK

2. William Harvey Research Institute, Queen Mary University of London, London, EC1M 6BQ, UK.

## Author Contributions

Conceptualization AMF

Data curation WC, SFA, AS, RT, AM, CD, SW, Genomics England

Investigation WC, IL, TB, CS, DO, HY, TL, LA, SHar, Sh, GB, DB, PC, NP, PVL, AMF

Writing WC, IL, PC, PVL, AMF

## Additional information

Refer to Web version for supplementary materials and methods.

