## Supplementary figures and stats for "A Genetic Model for Central Chondrosarcoma Evolution Correlates with Patient Outcome"

**Supplementary Materials**

***Supplementary figures***

Supp. Figure 1 – Structural changes near the *TERT* gene

Supp. Figure 2 – Analysis of drivers dNdS

Supp. Figure 3 – Methylation array clustering outline

Supp. Figure 4 – Genome doubling classifier

Supp. Figure 5 – Outline of mutational channels used for signature analysis

Supp. Figure 6 – Outline of DD CS statistics

Supp. Figure 7 – Clock like mutational signatures

Supp. Figure 8 – Full outcome data, inc *TERT* methylation

***Supplementary Notes***

Note 1 – Visual confirmation of *IDH1*, *IDH2*, and *TERT* hotspot mutations

Note 2 – Flow cytometry confirmatory analysis of ploidy

Note 3 – Digital PCR parameters and design tables

***Supplementary tables***

Supp. Table 1 – Overview of Genomics England 100KGP cases

Supp. Table 2 – Overview of clinical data and digital PCR results

Supp. Table 3 – Outline of driver calls in 100KGP data

Supp. Table 4 – Outline of methylation driver calls

*Supplementary Figure 1 – Structural changes near the TERT gene*

A: WGS_16

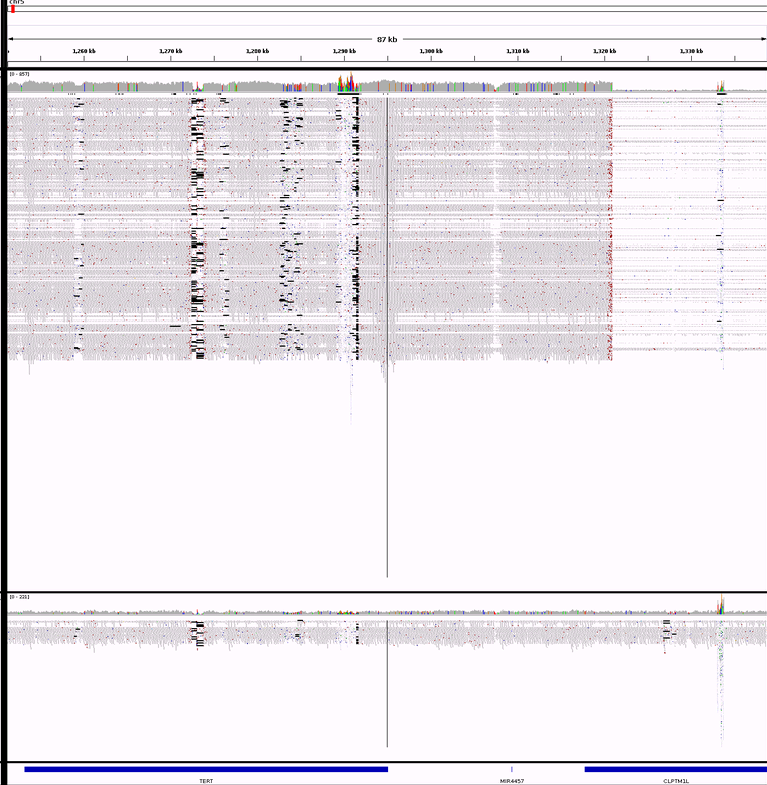

chr5:1320965

B: WGS_19

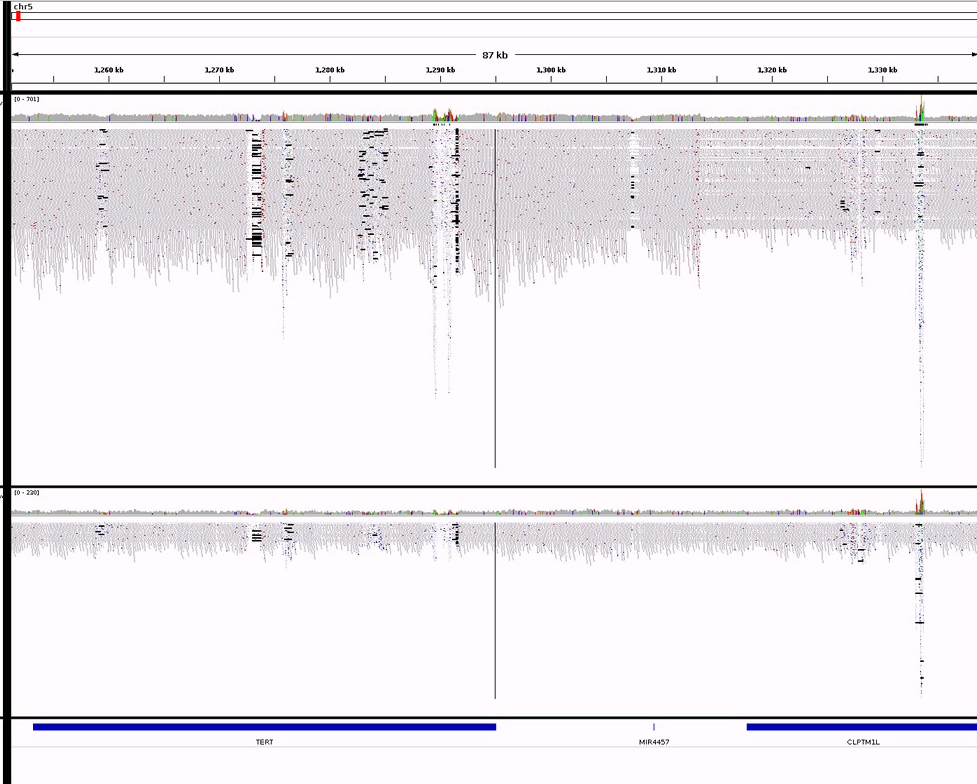

chr5:1313467

C: WGS_21

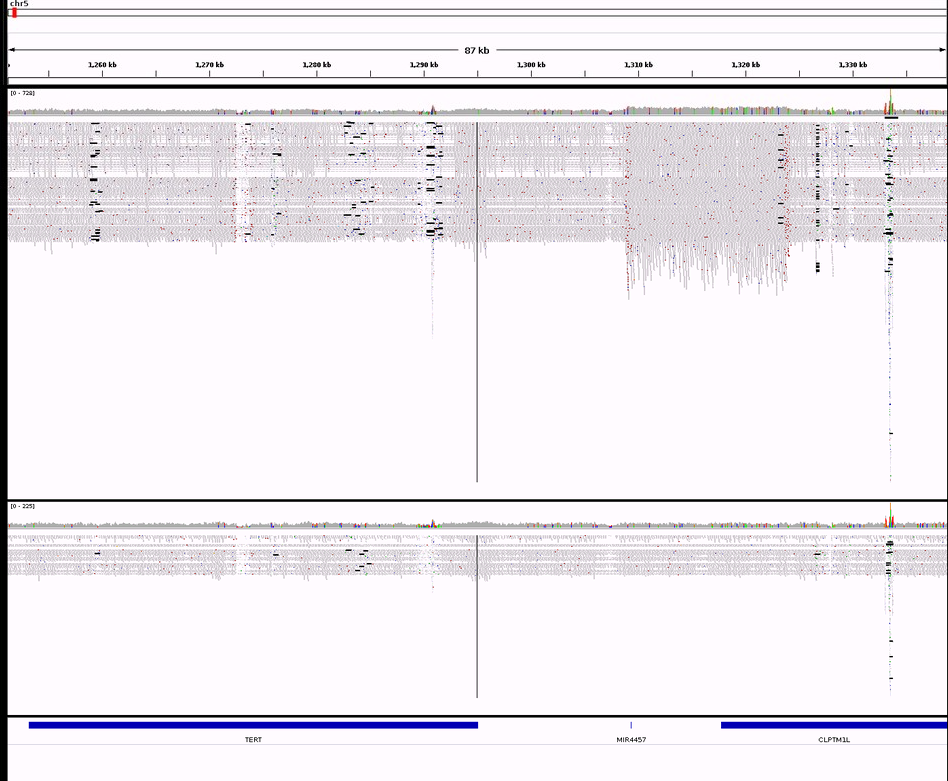

chr5:1297860

D: WGS_68

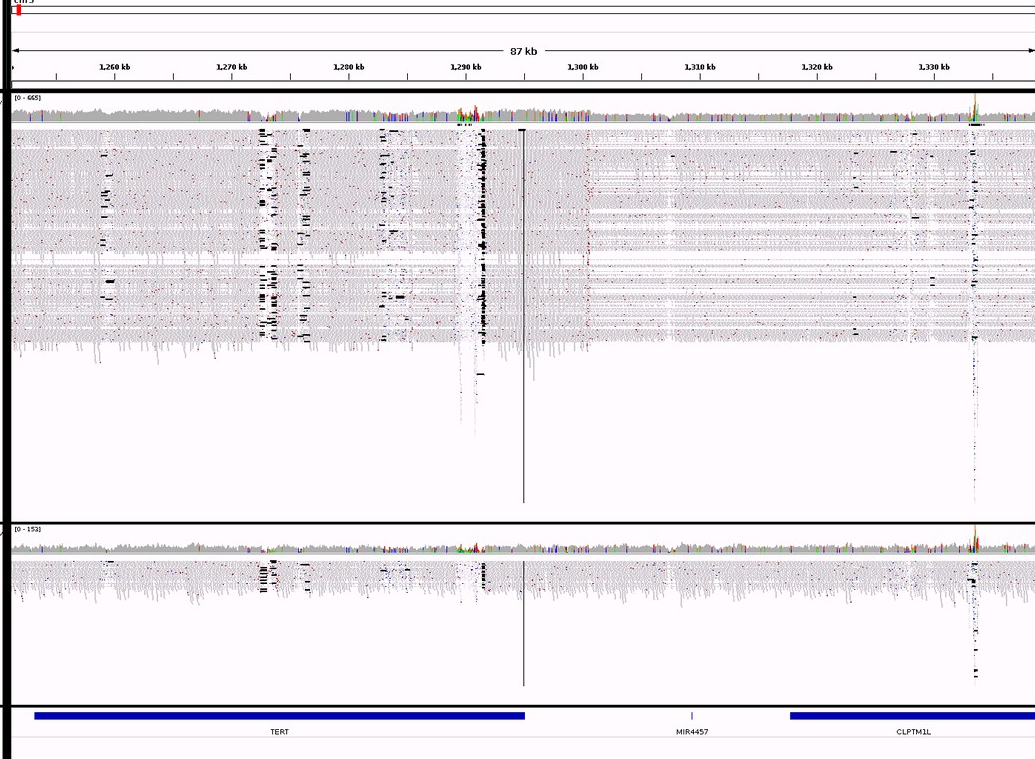

chr5:1300678

IGV screenshots of the *TERT* gene and promoter loci. Images **A-D** are reads associated with tumour genomes (shown above) and paired normal (below). Aberrations highlighted by red arrow and boxes contain chromosome start positions. Note: *TERT* gene positions are indicated by the leftmost, lower blue rectangle. The duplication of Plot C (WGS_21) overlaps the single chromothripsis event identified in this cohort. The start and stop position for this event was chr5:14757-46420988 and it contained 32 breakpoints, encompassing this and 18 other SV events.

*Supplementary Figure 2 – Analysis of drivers dNdS*

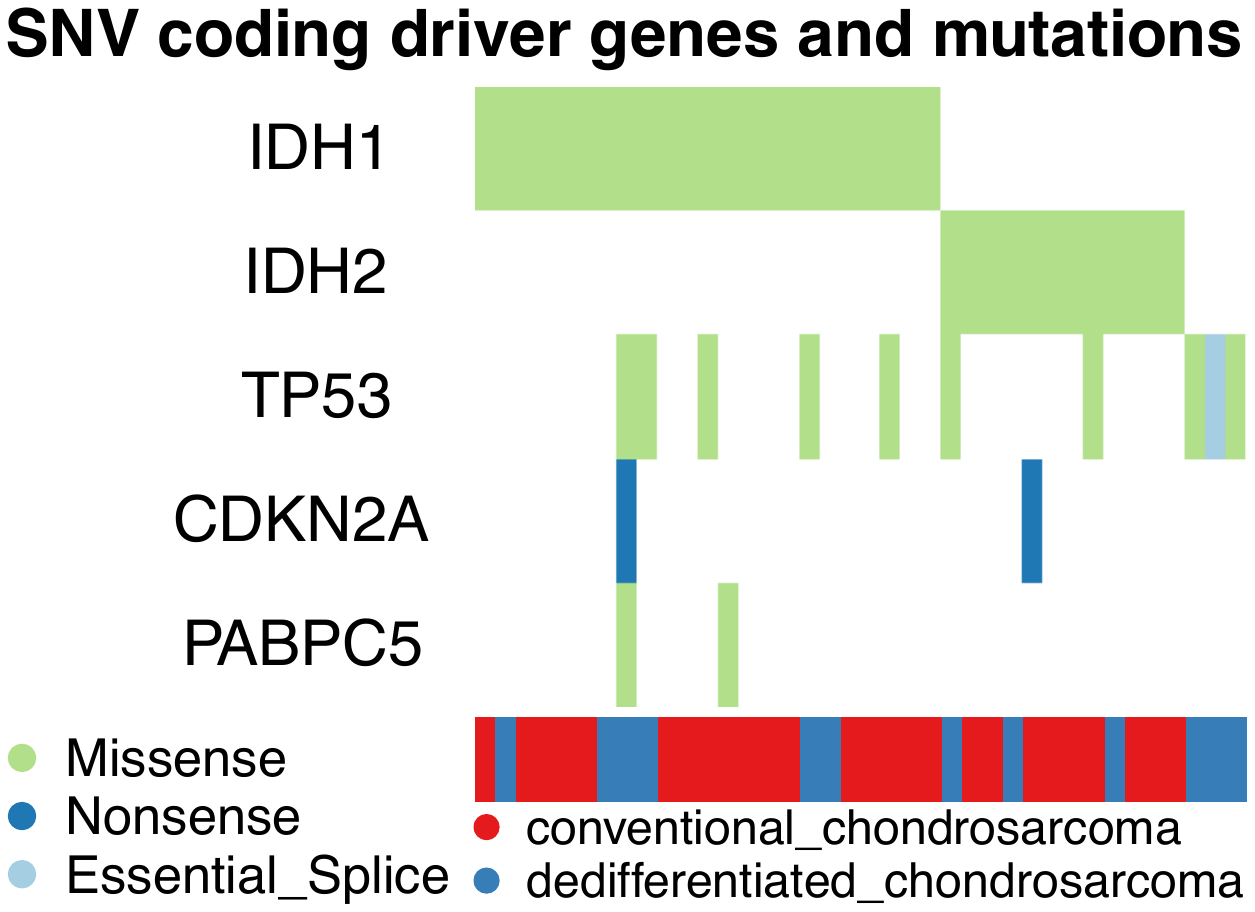

Tile plot of genes significantly altered according to dNdS analysis, across 38 chondrosarcomas with these mutations.

*Supplementary Figure 3 – Methylation array clustering outline*

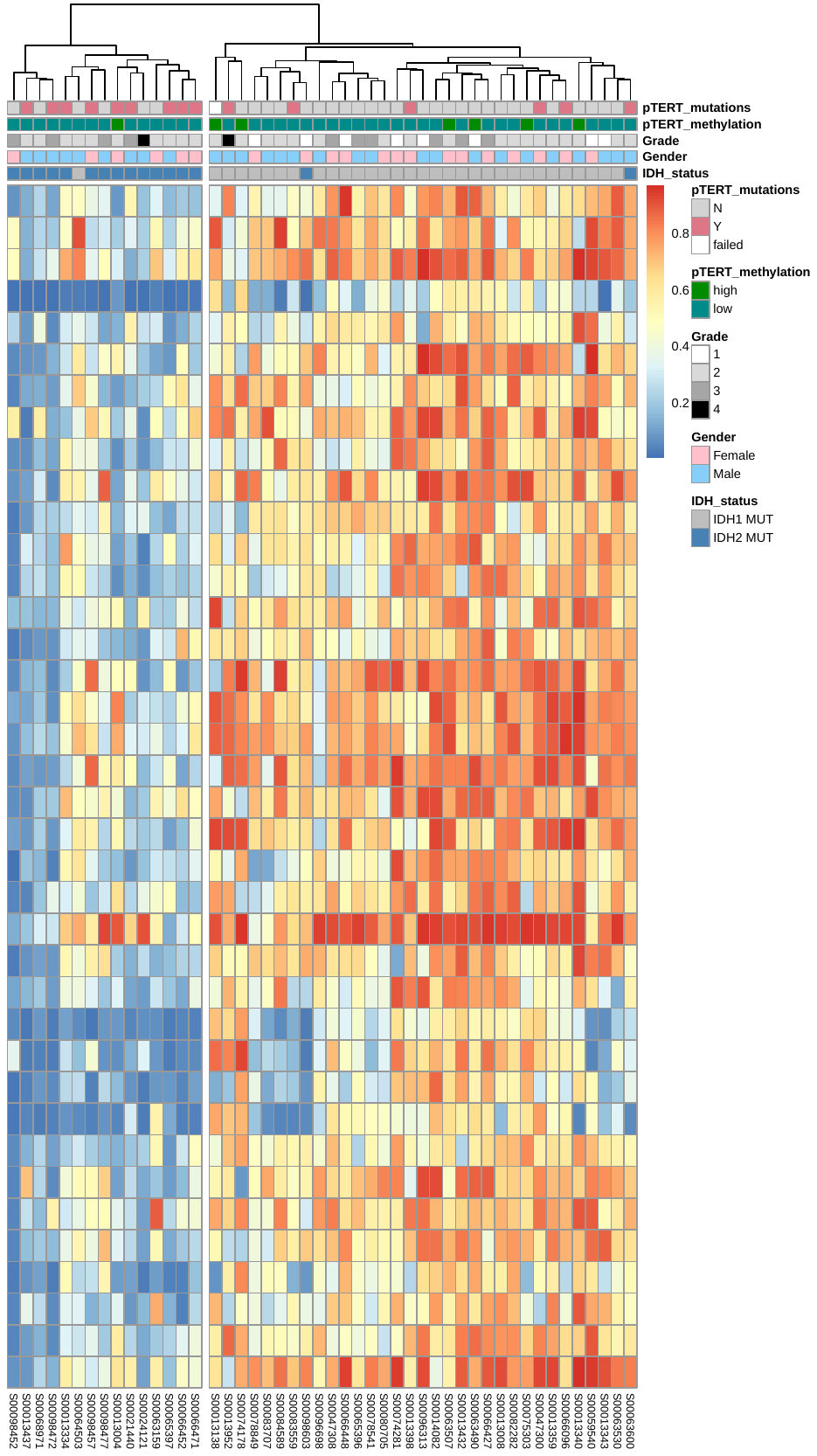

Supervised clustering plotted using the Pheatmap R package. The 48 most differential methylated probes between *IDH1* and *IDH2* mutant cases are shown. Left clade shows *IDH2*-mutant tumour methylation profiles, right shows those related to *IDH1*.

*Supplementary Figure 4 – Genome doubling and haploidisation classifier*

A	B

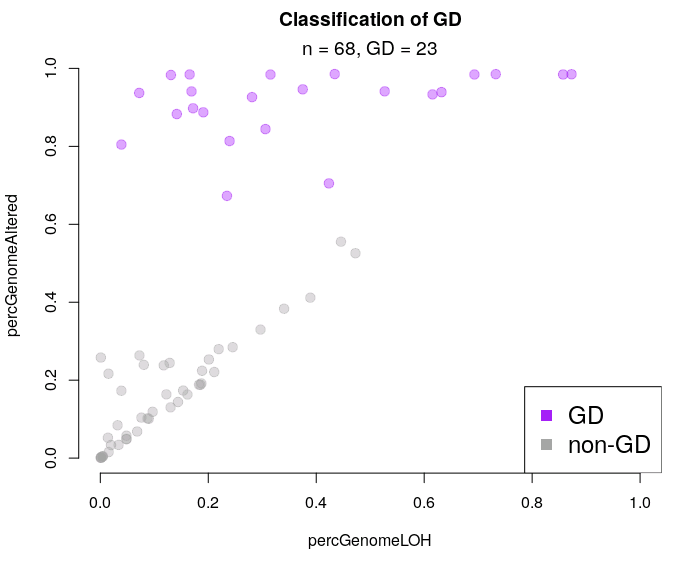

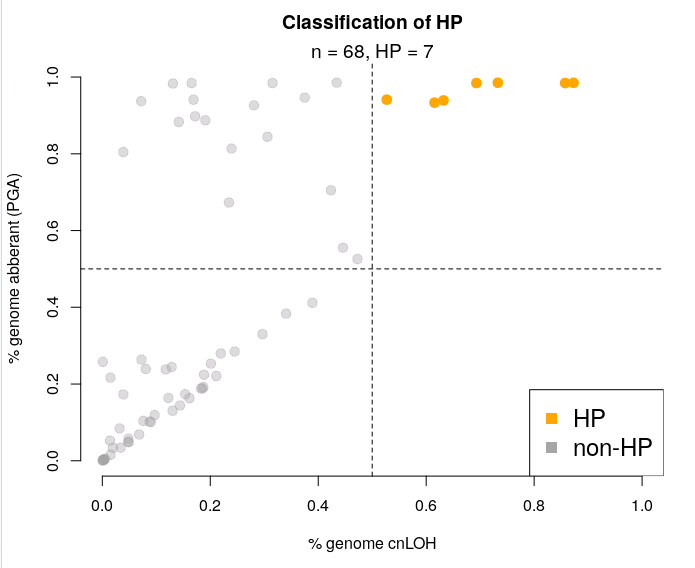

C	D

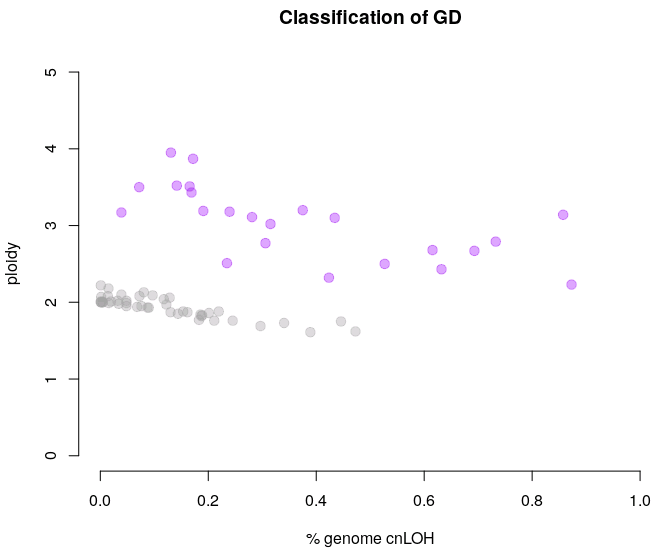

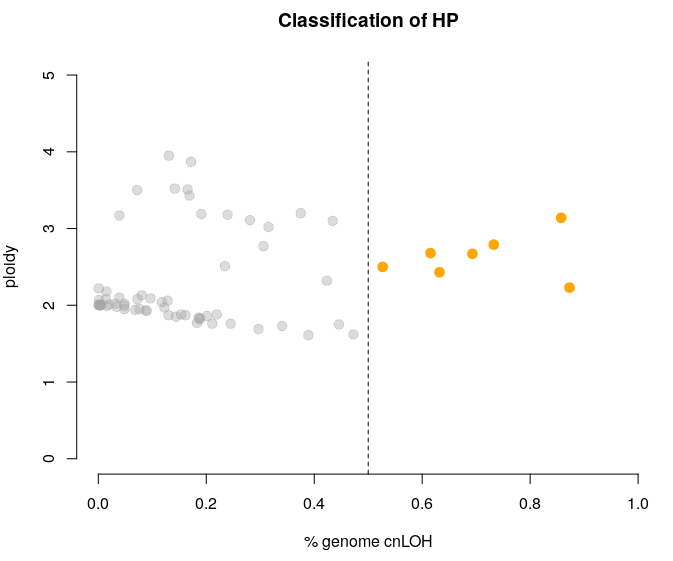

Results of genome doubling classified based on the R Mclust function. Scatter plots A and B show the classification results (**A**: genome doubling in purple, **B**: haploidy cases in orange). Genome doubled cases generally have a high percentage of genome aberrant (PGA > 80%, y-axis). Haploidy were classified as those with than 50% of their genome at cnLOH state (x-axis). Plots **C-D** show same data but with ploidy on y-axis.

*Supplementary Figure 5 – Outline of mutational channels used for signature analysis*

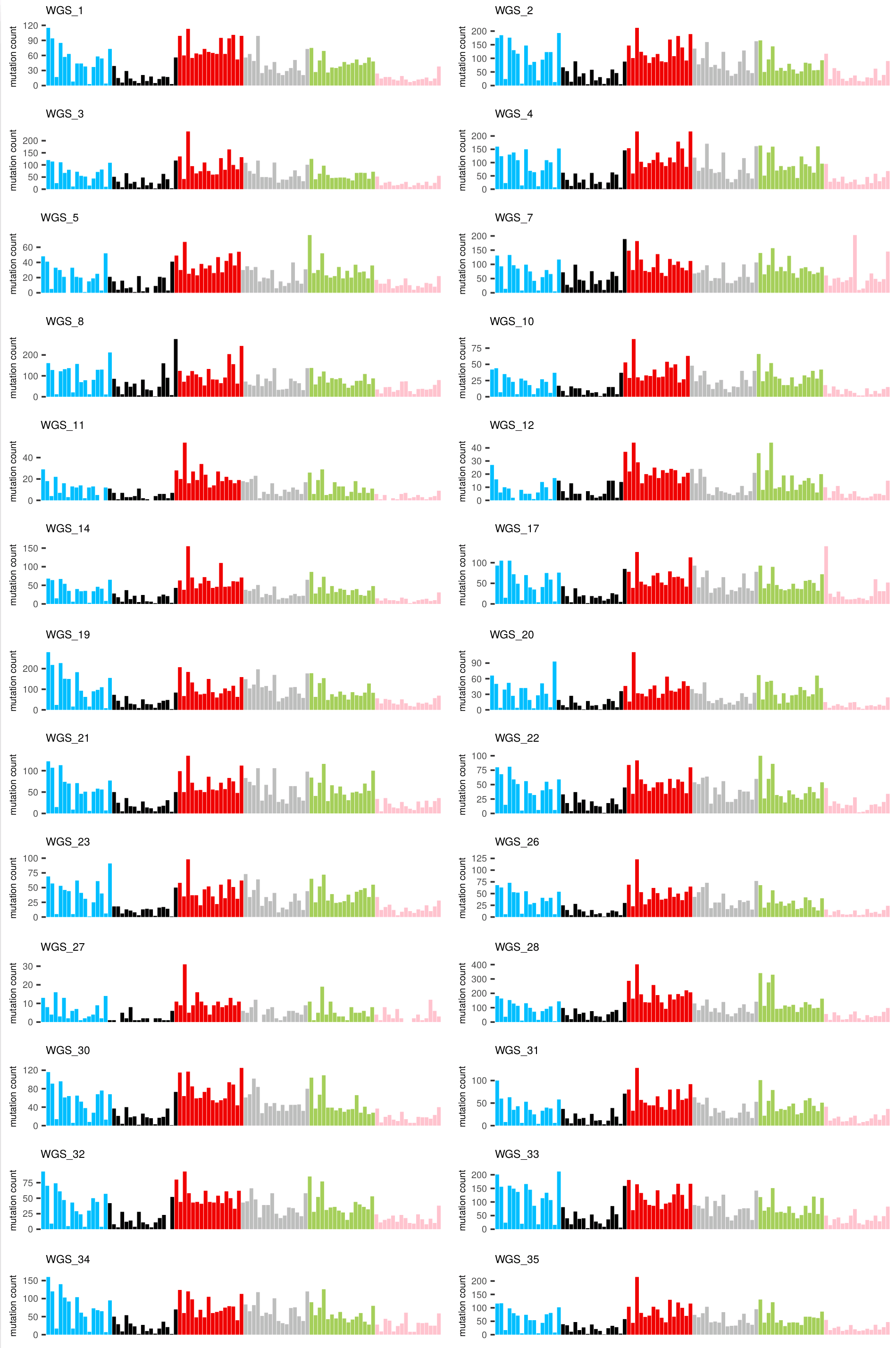

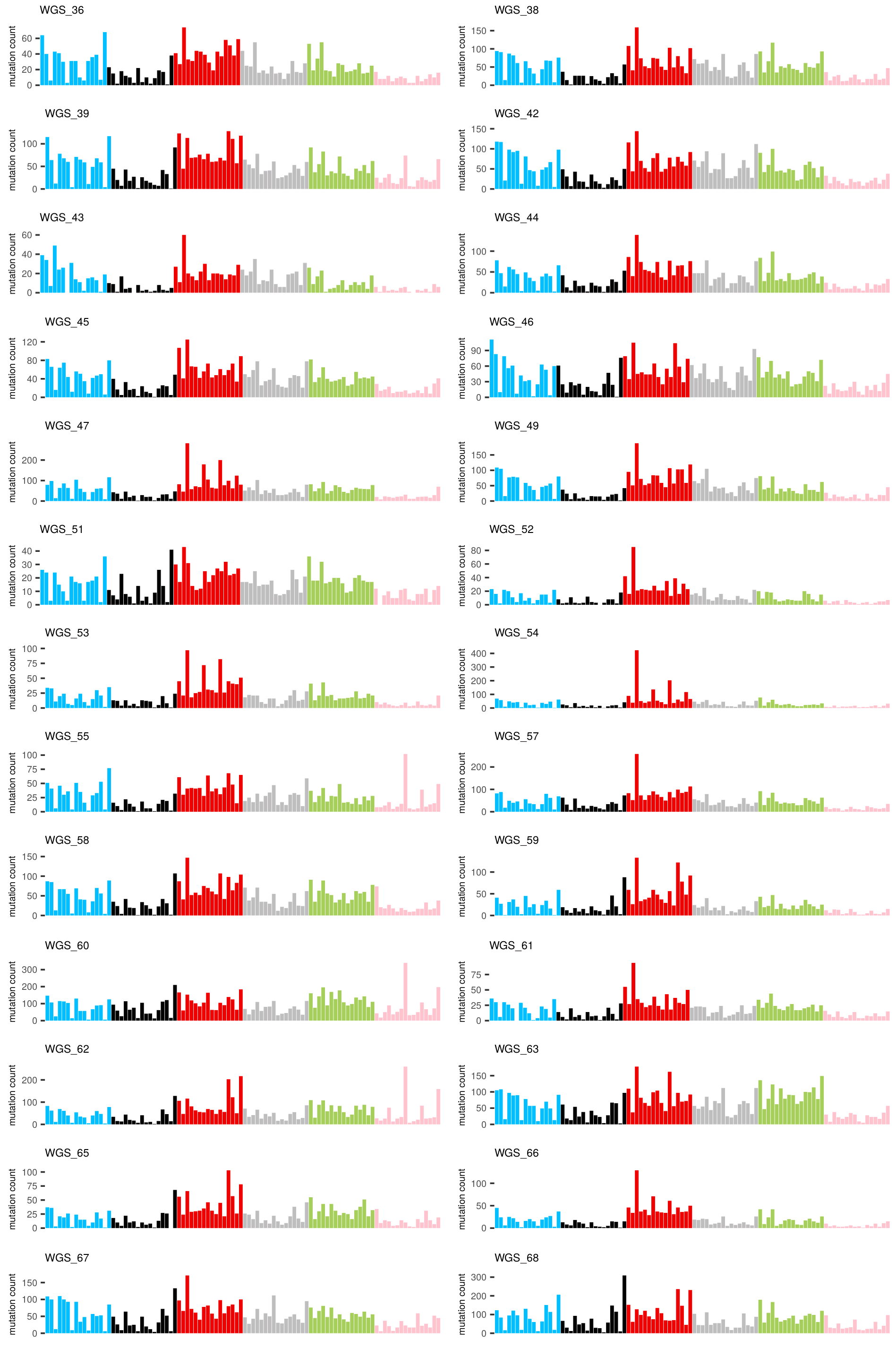

Bar plots of 96 channel 100KGP cases WGS_1 - 68. Y-axis shows the single nucleotide variant counts for each base context. X-axis shows the mutational context following the conventional order of C>A, C>G, C>T, T>A, T>C, and T>G. These data were used as input for the analysis utilising SigProfiler.

*Supplementary Figure 6 – Outline of DD CS statistics*

**A	B**

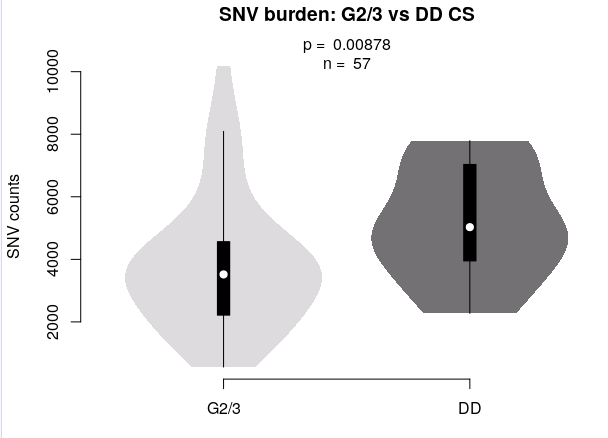

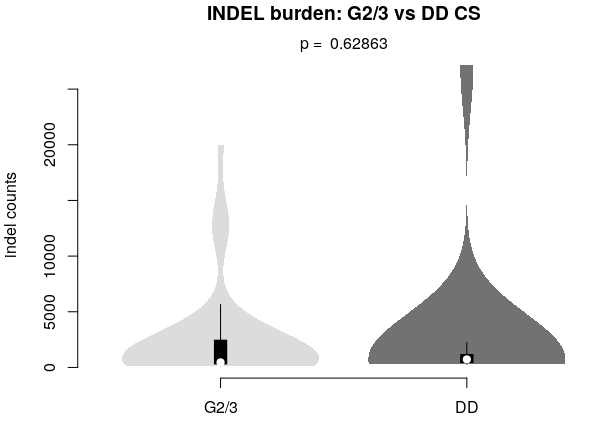

**C	D**

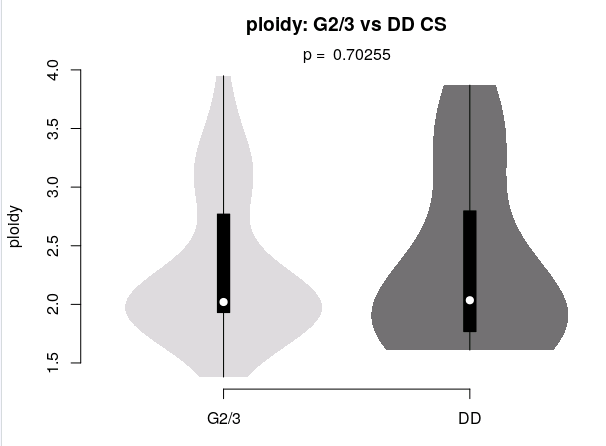

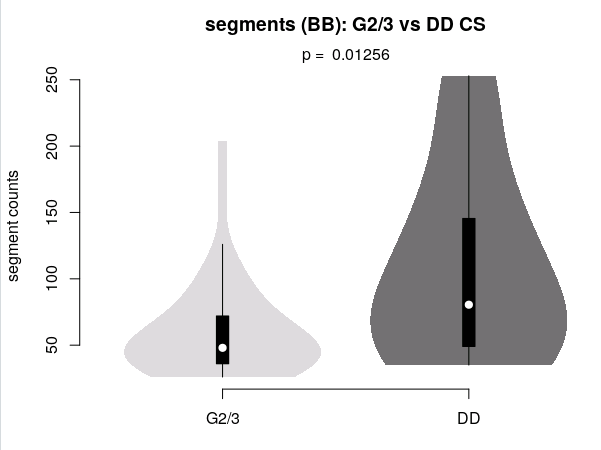

**E	F**

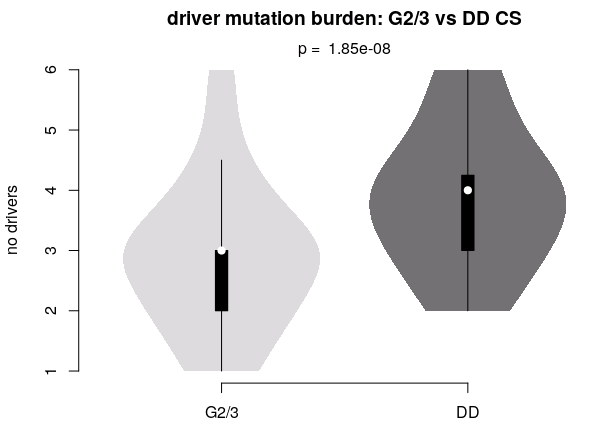

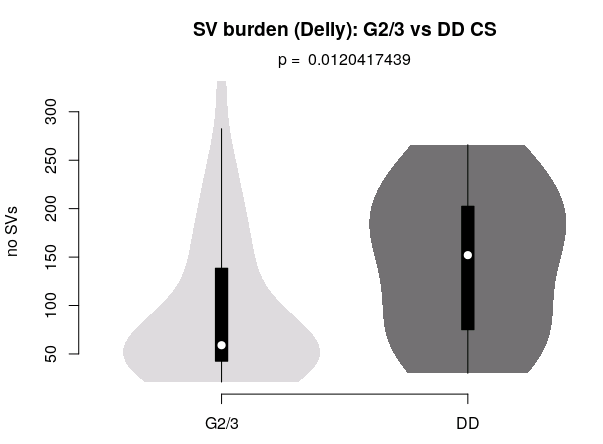

Summary statistics and violin plots for comparisons of G2/3 and DD CS cases, using: SNV burden (**A**), Indel burden (**B**), ploidy (**C**), number of Battenberg derived segments (**D**), total driver burden, defined by figure 1D in the main text (**E**), and SV burden derived from the Delly calls (**F**). Statistically significant results are highlighted by red boxes.

*Supplementary Figure 7 – Clock like mutational signatures*

**A	B**

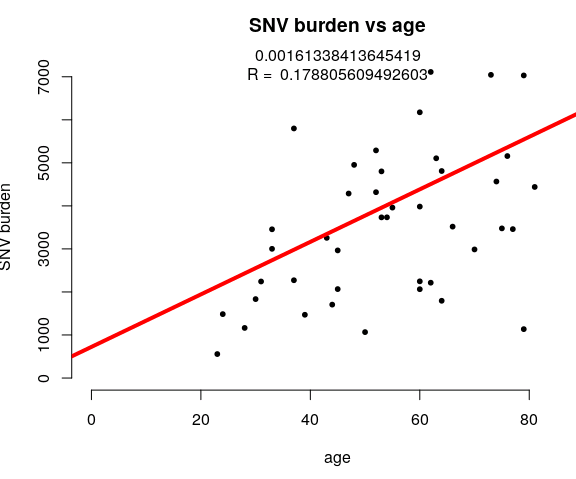

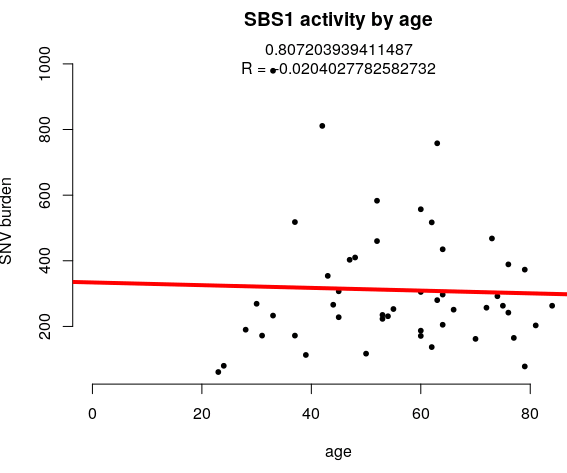

**C	D**

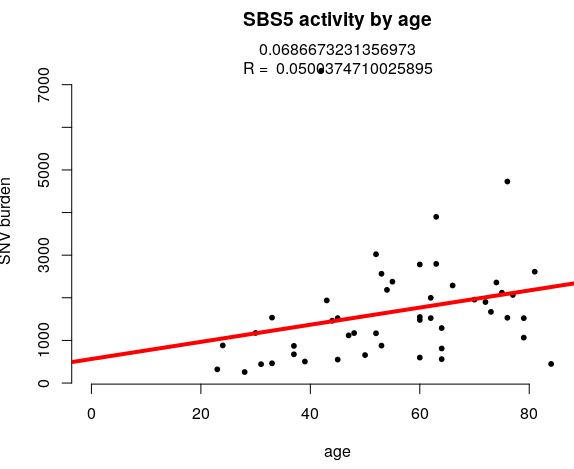

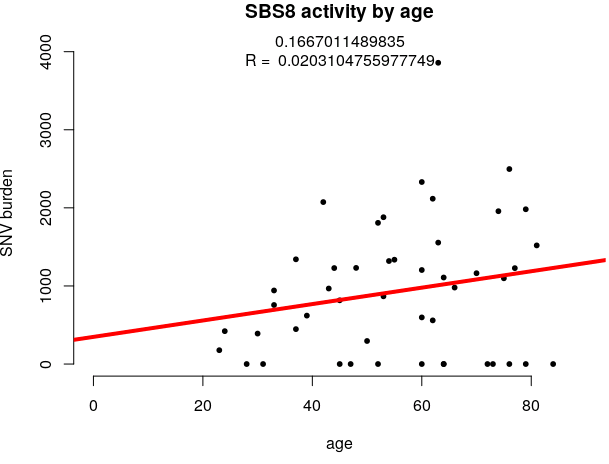

**E	F**

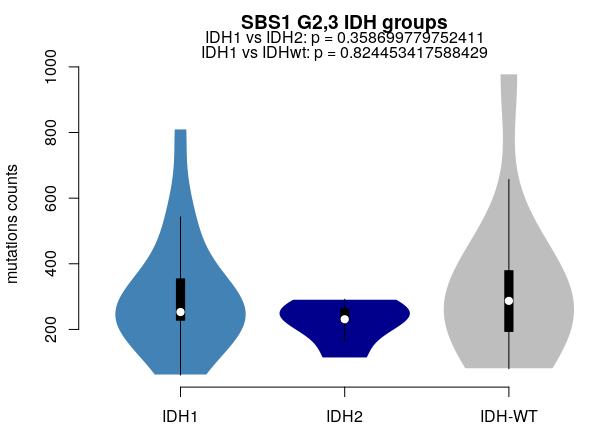

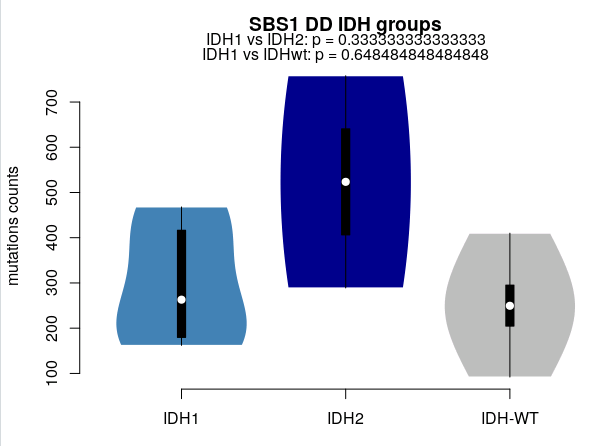

**G	H**

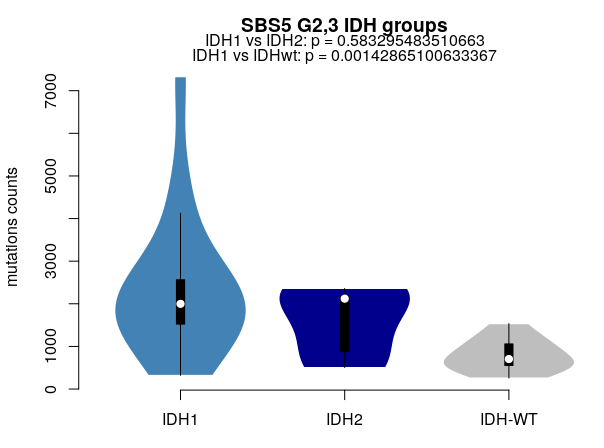

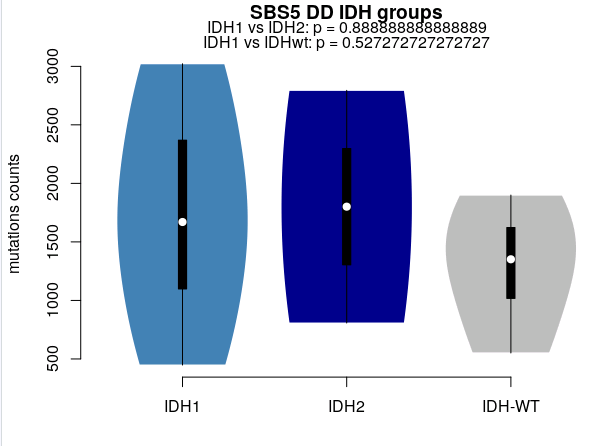

**I	J**

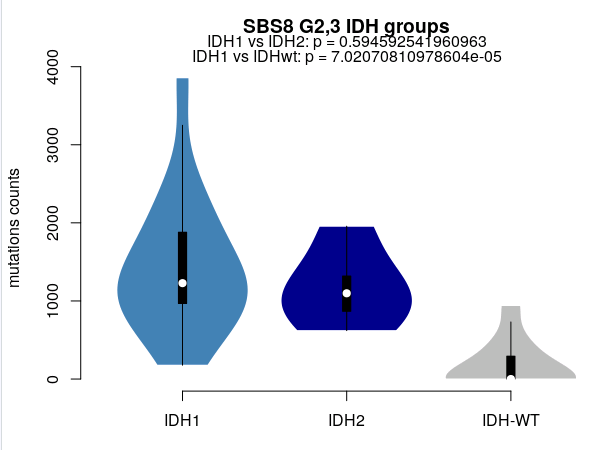

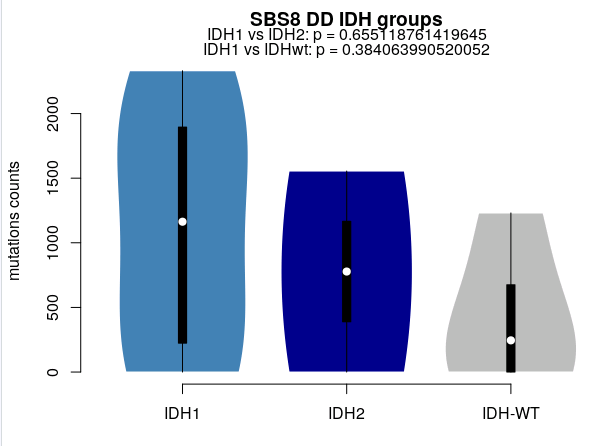

**K	L**

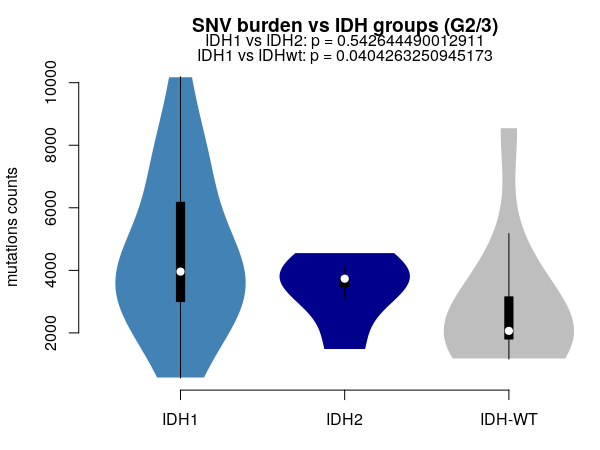

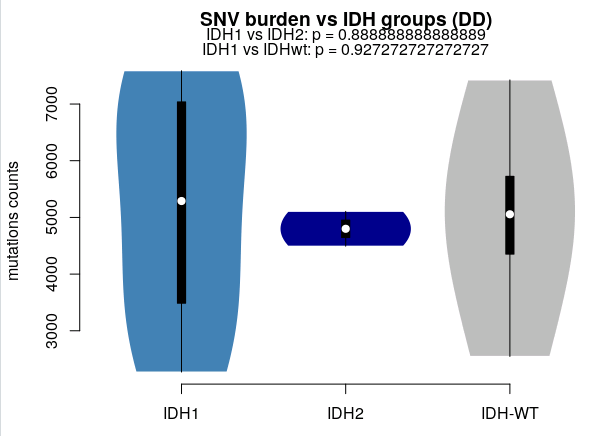

Summary of mutational clock analysis. Scatter plots **A-D** demonstrate relationships (or lack thereof) between signature counts (SBS1, SBS5, and SBS8), and total SNV burden, against patient age at diagnosis. Red line indicates linear regression. R^2^ and p values shown above. Violin plots E-L are grouped by IDH status and show significant differences in *IDH1*-IDHwt / *IDH1*-*IDH2*, by each signature activity in G2/3 and DD CS cases. Significant results indicated by red lines.

*Supplementary Figure 8 – Outcome data*

**A** *TERT* methylation: HR 3.4, p = 0.02	**B.** *TERT* meth / mut: HR 3.2, p = 0.03

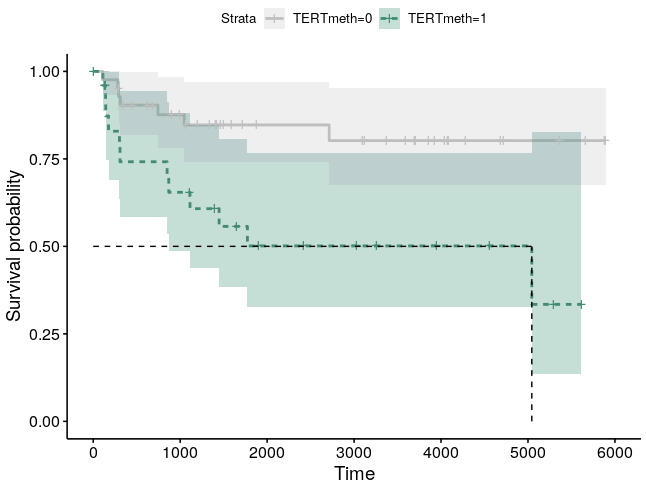

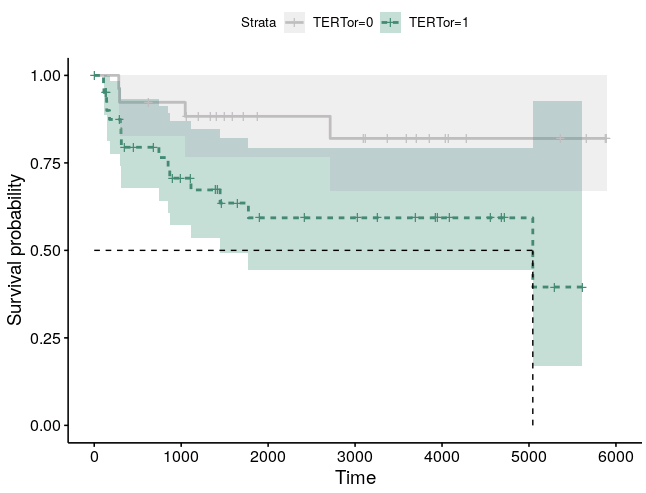

**C**	All clinical factors (*IDH1* and *IDH2* separate)

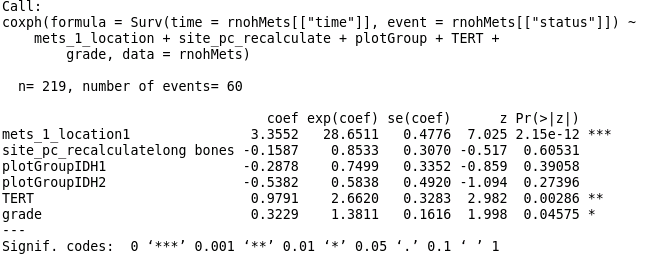

**D**	All clinical factors (*IDH1* and *IDH2* merged)

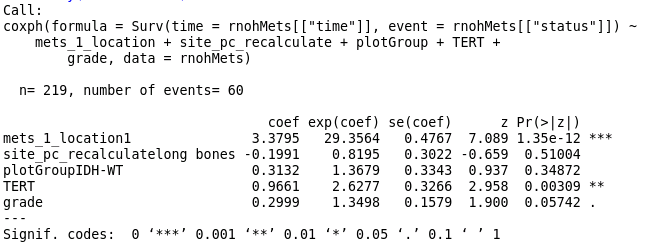

Kaplan-Maier plots (**A**) demonstrate the association of outcome and *TERT* methylations status. Plot (**B)** shows grouped outcome of canonical *TERT* promoter mutations and/or methylation status. Results of cox proportional hazard model for all established clinical parameters (screen print **C**), including: presence of metastasis/local recurrence (*mets_1_location*), anatomical location (axial or long bones: *site_pc_recalculate*), IDH status (plot group), canonical *TERT* promoter mutation status, and grade. Anatomical site and IDH groups are non-significant in this context, though IDH1/2 status is significant in isolation (see main Figure 3). The same test but with *IDH1* and *IDH2* status combined is shown in screen print **D**.

***Supplementary tables***

*Supplementary Table 1 – Outline of 350 chondrosarcoma cases*

*Supplementary Table 2 – Genes in SVs table*

***Supplementary Notes***

*Note 1 – Visual confirmation of IDH1, IDH2, and TERT hotspot mutations*

WGS_1: *IDH1* R132G

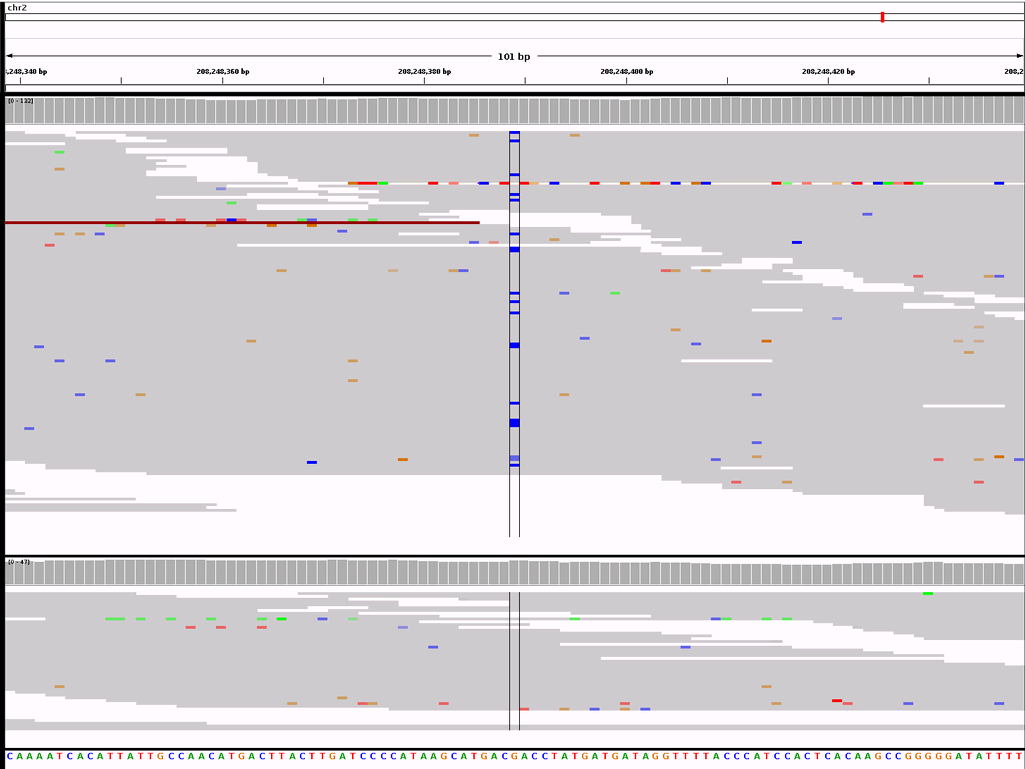

WGS_12: *IDH1* R132C

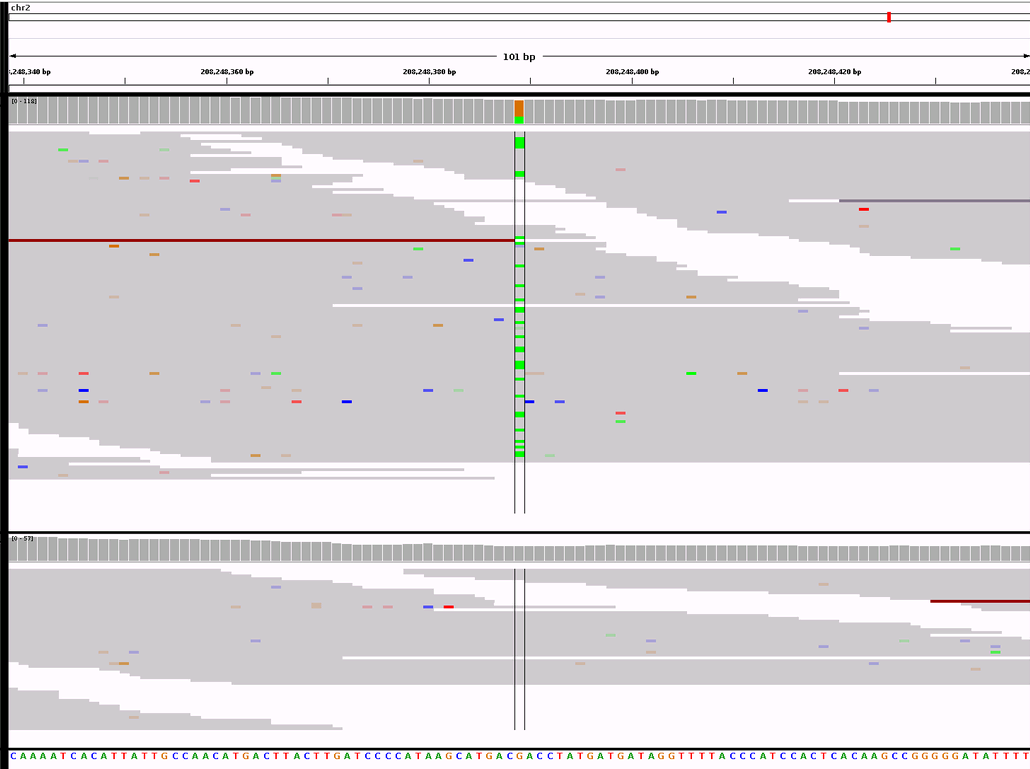

WGS_43: *IDH2* R172S

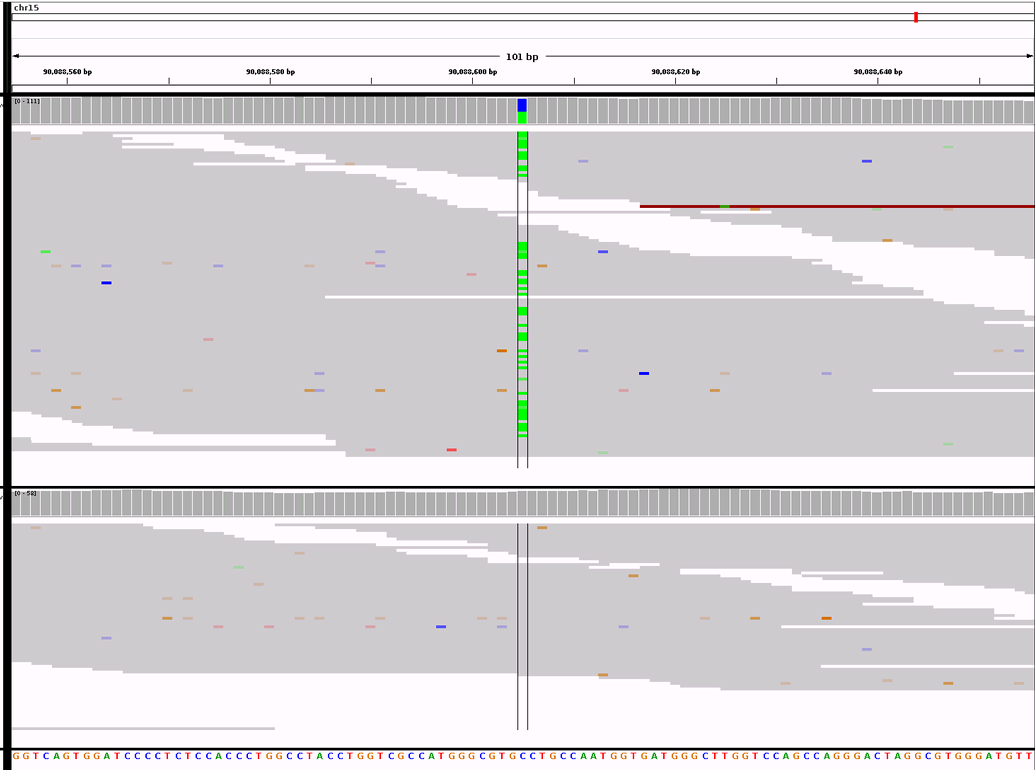

WGS_53: *IDHwt* (ddPCR result *IDH1*)

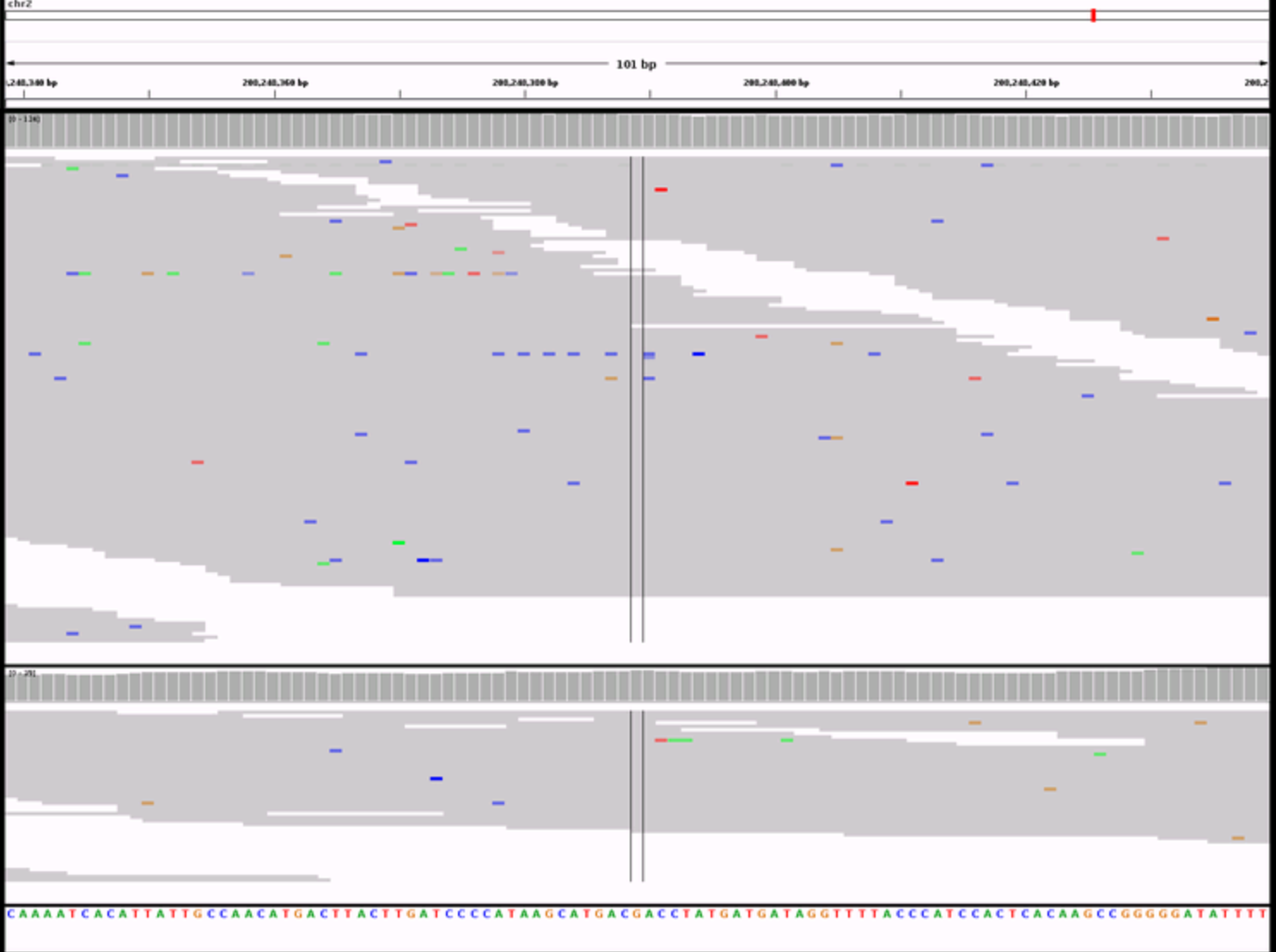

WGS_5: *TERT* (1295113 G>A)

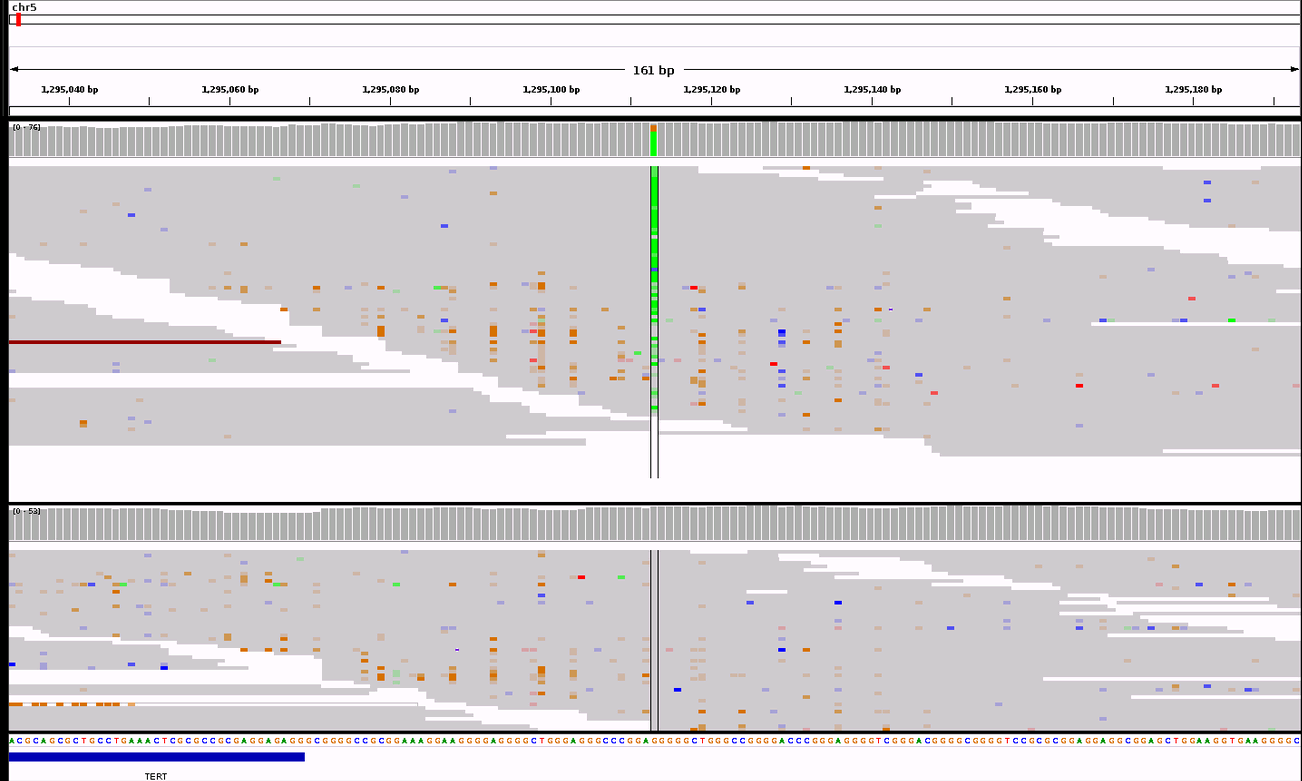

WGS_6: *TERT* (1295113 G>A)

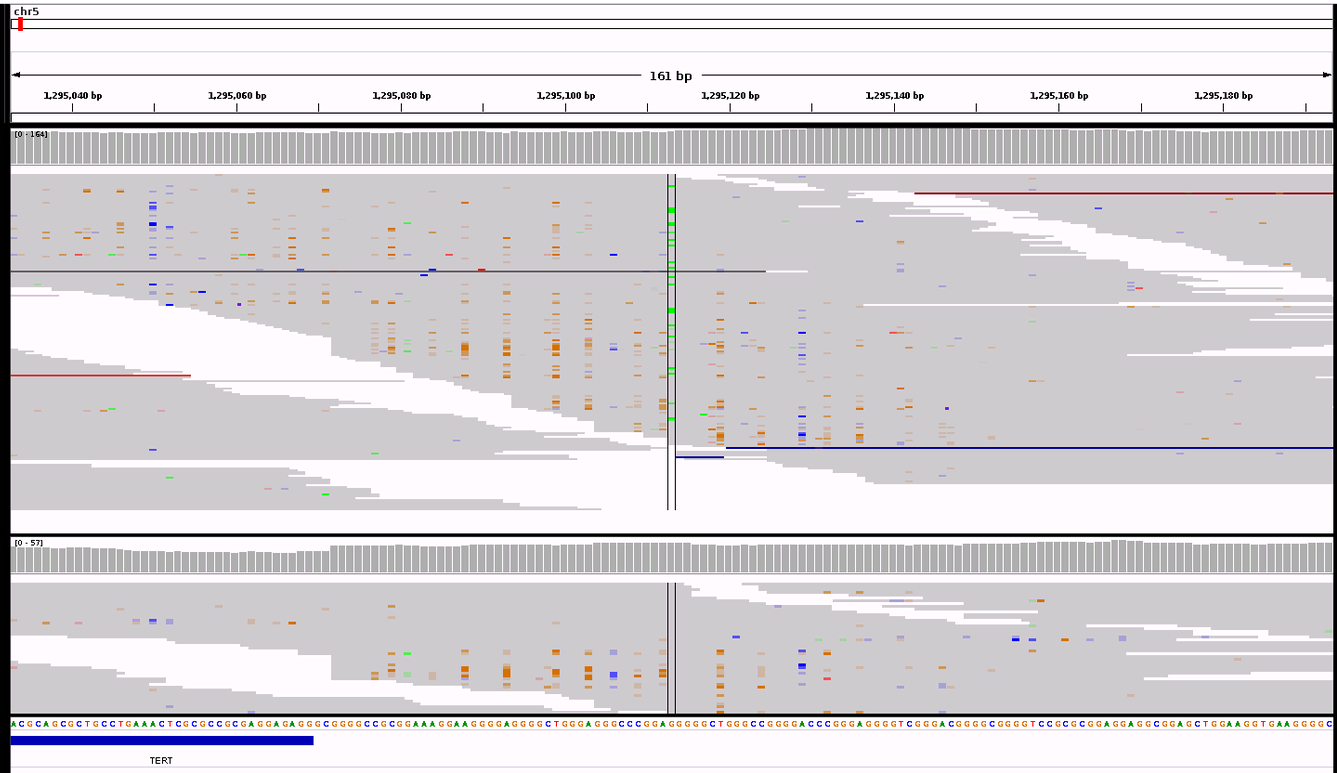

As part of the validation process we visually examined all mutations in *IDH1*, *IDH2*, and *TERT* called by Strelka in the 100KGP data. We used IGV for this purpose and confirmed all but one of the mutations in 60 of cases using digital PCR (WGS_53, see **Online Methods**). Above are 5 representative IGV screenshots.

*Note 2 – Flow cytometry confirmatory analysis of ploidy*

**A:** WGS_1

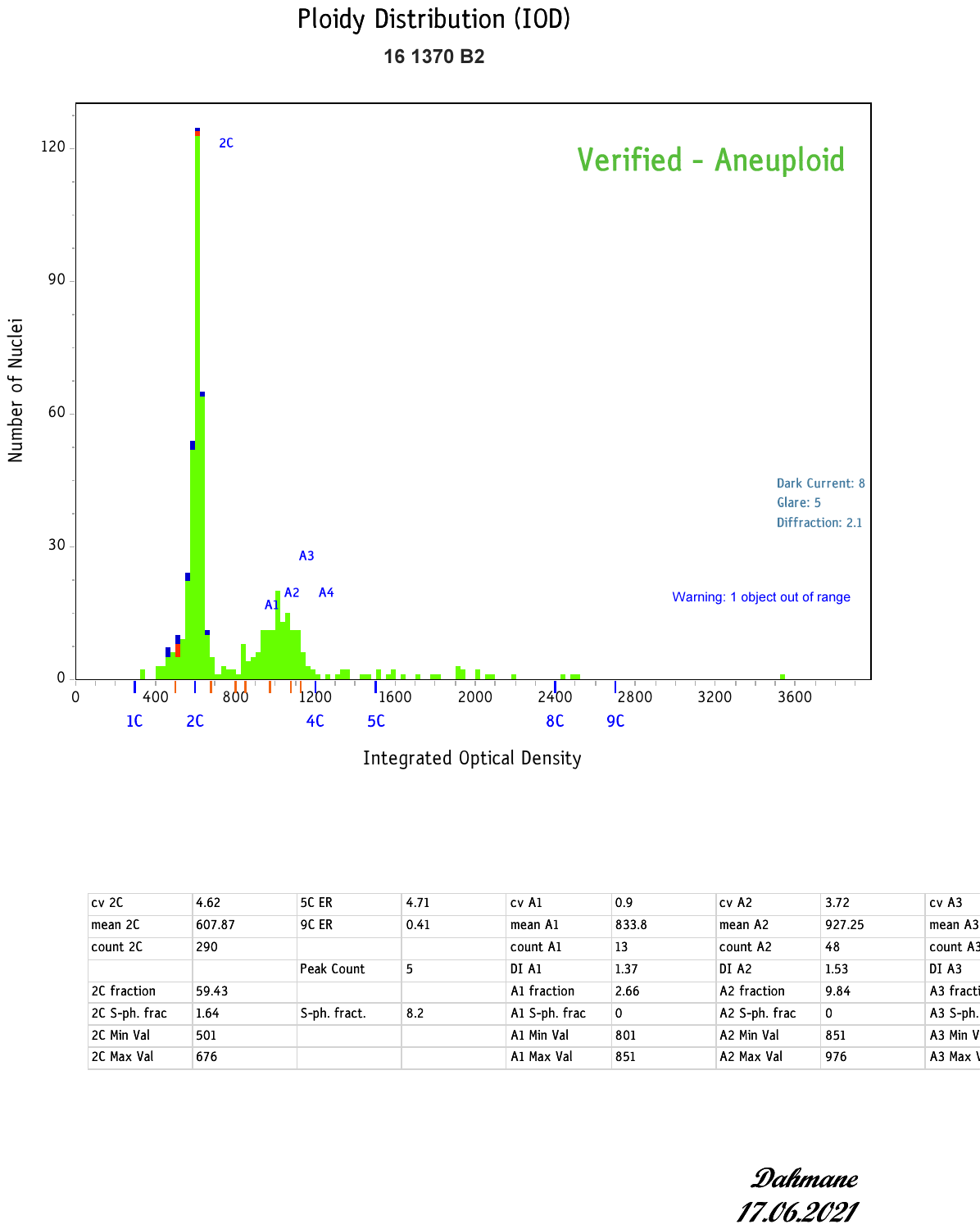

**B:** WGS_2

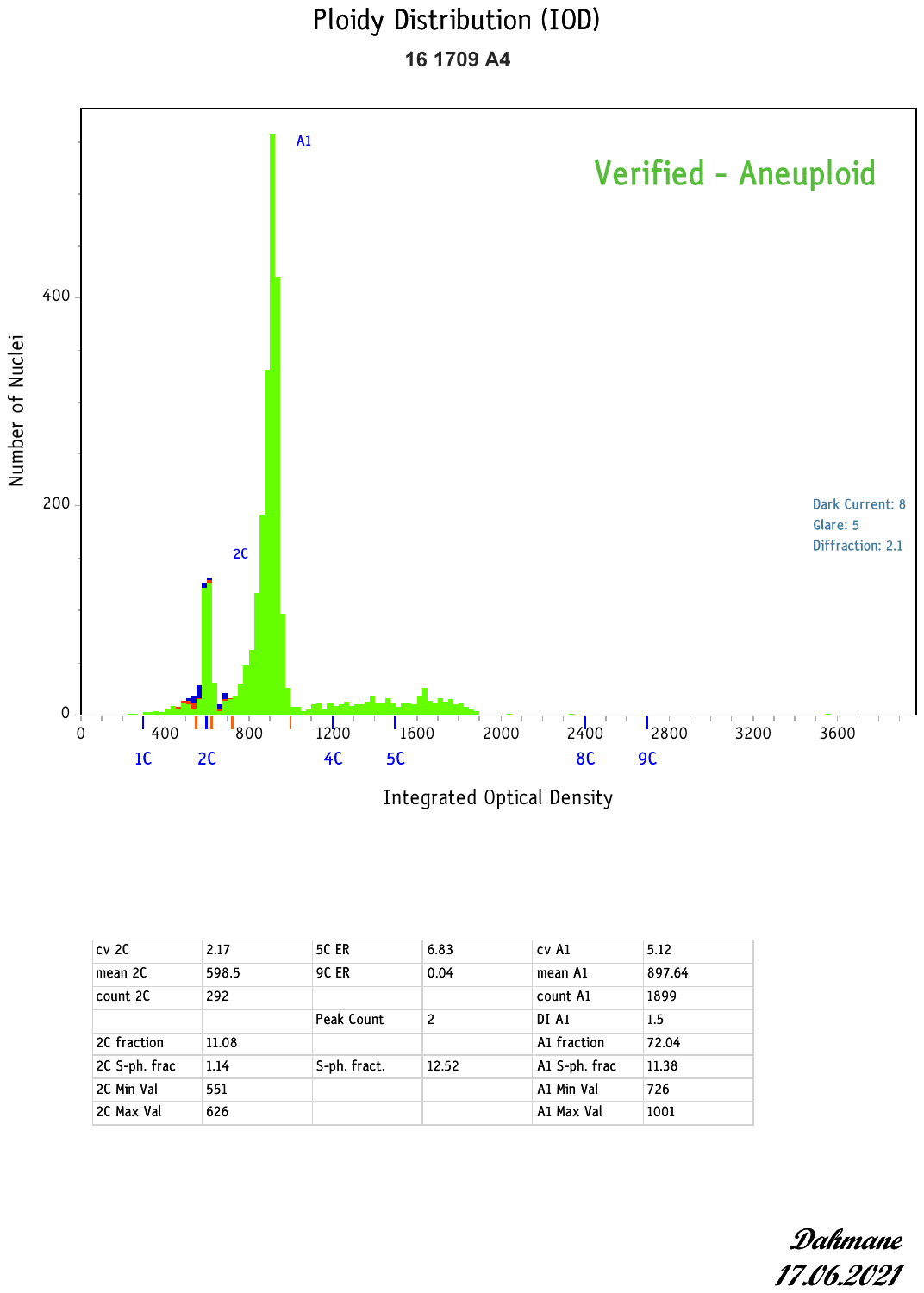

**C:** WGS_3

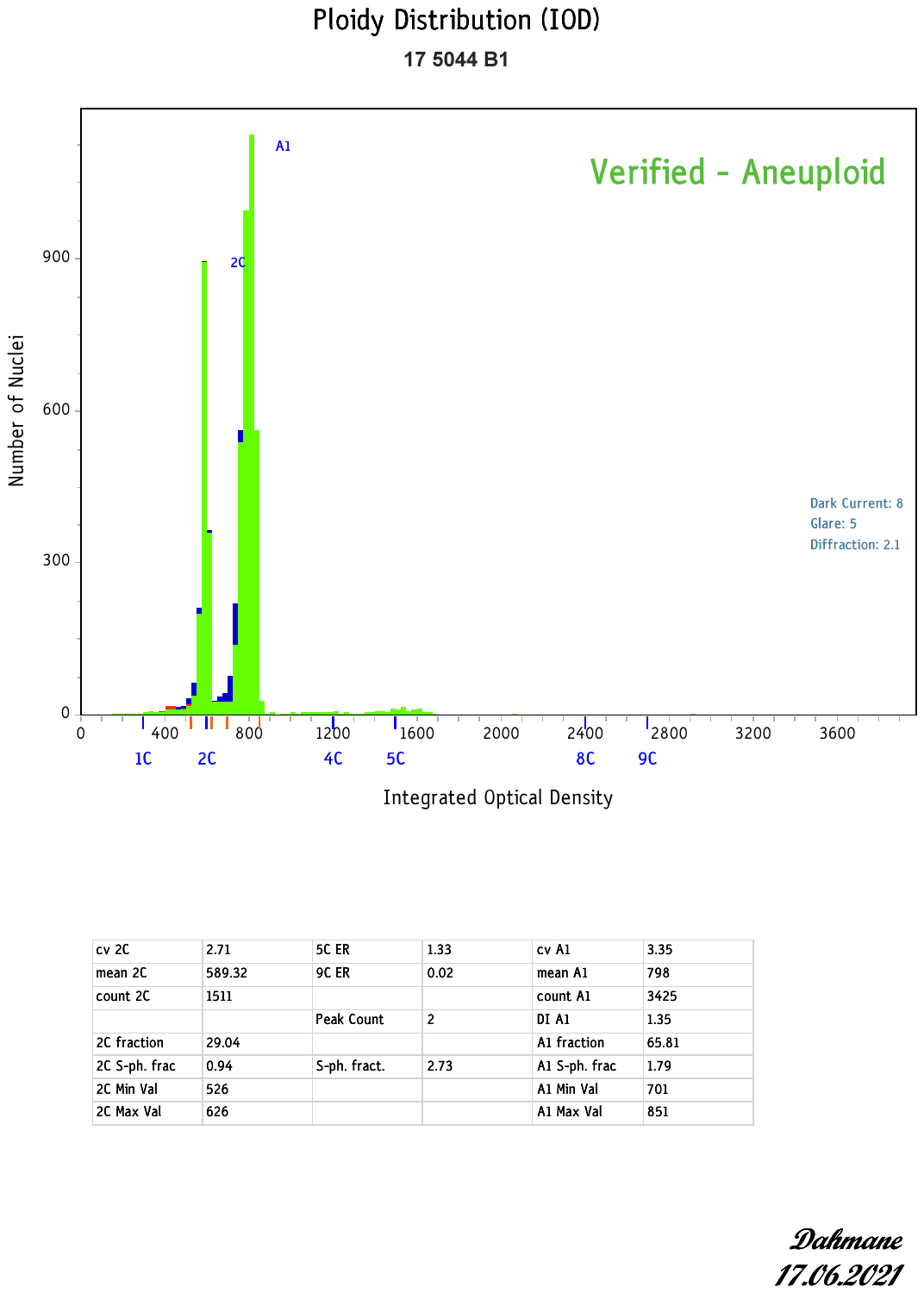

**D:** WGS_4

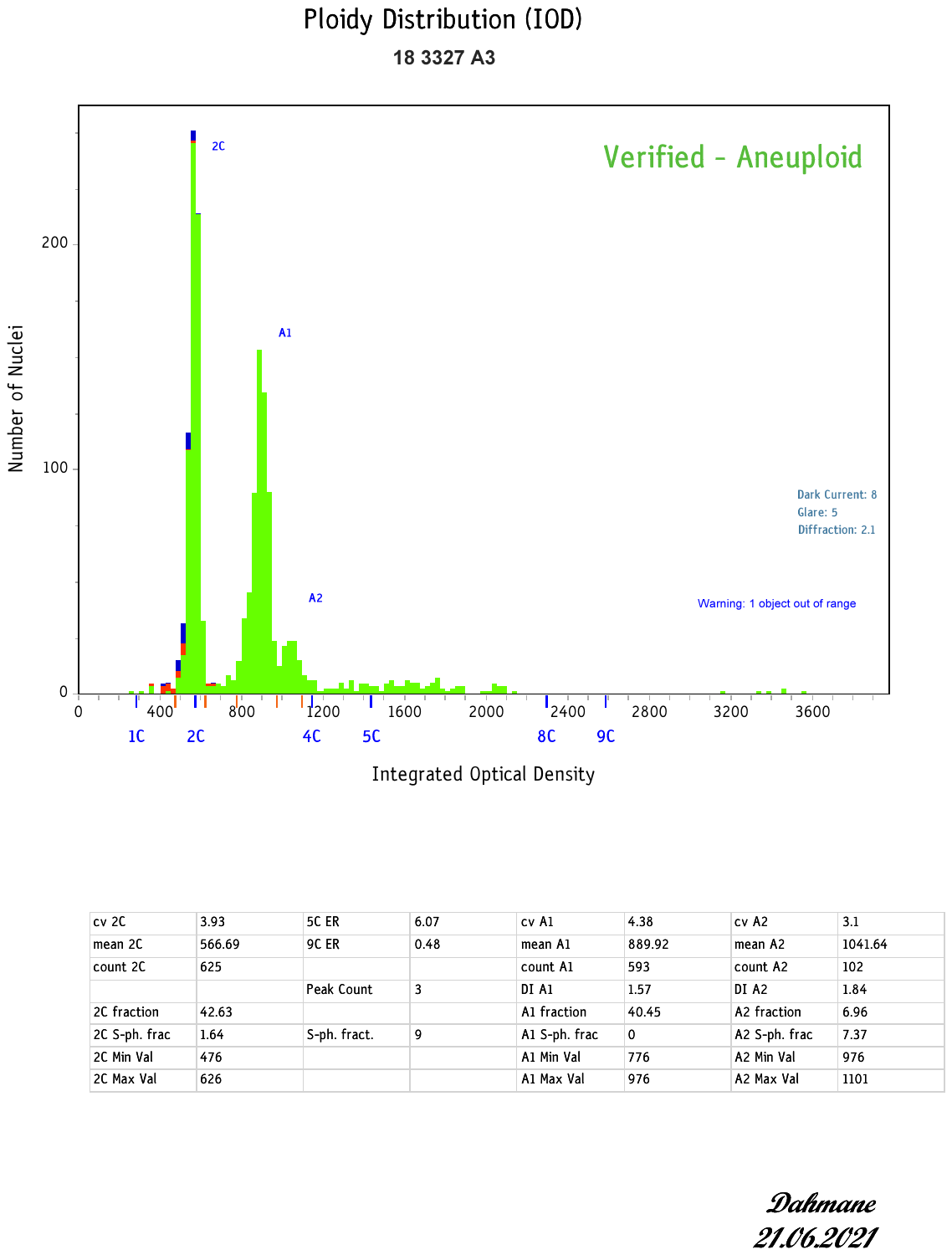

**E:** WGS_5

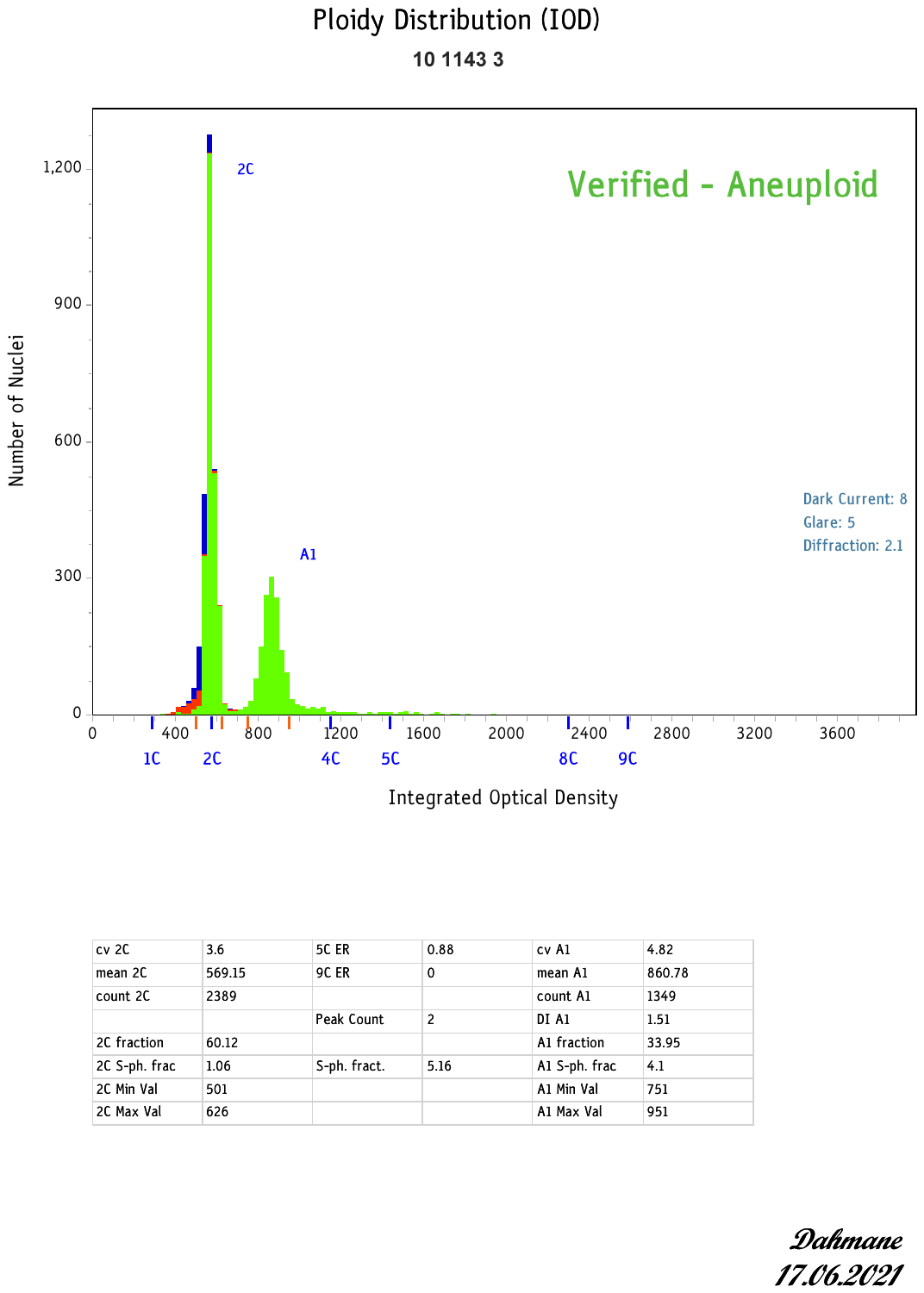

**F:** WGS_6

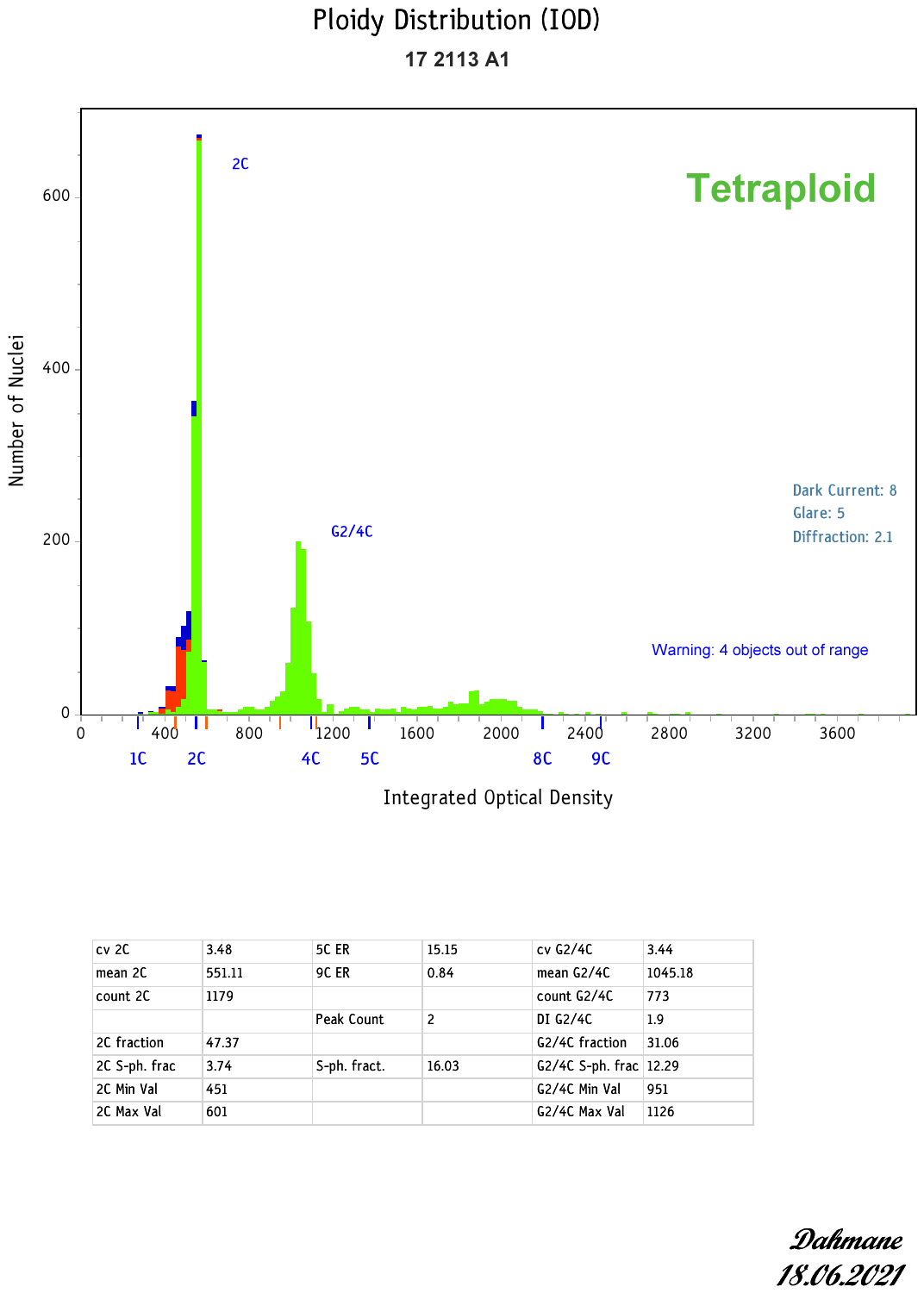

**G:** WGS_7

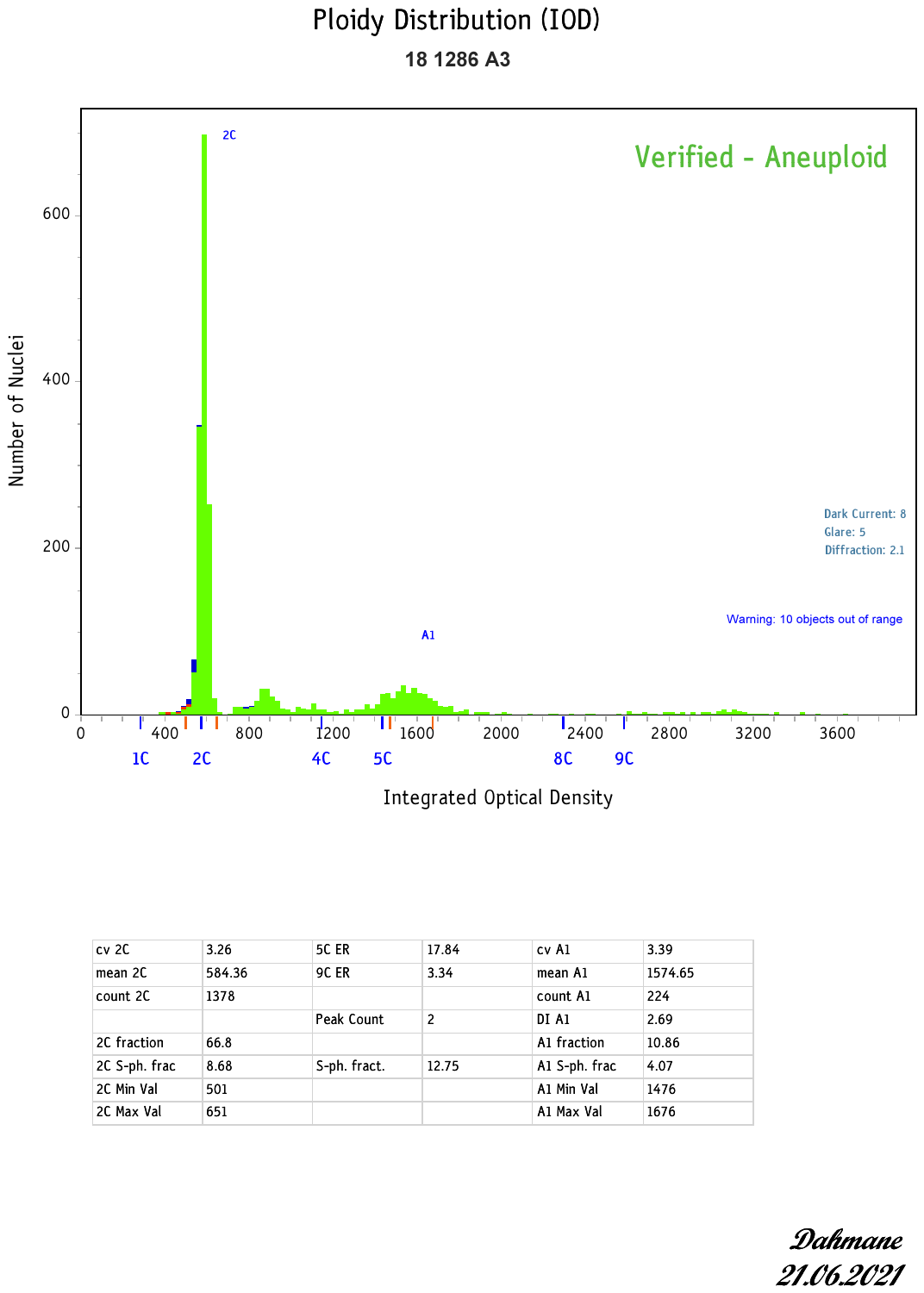

**H:** WGS_51

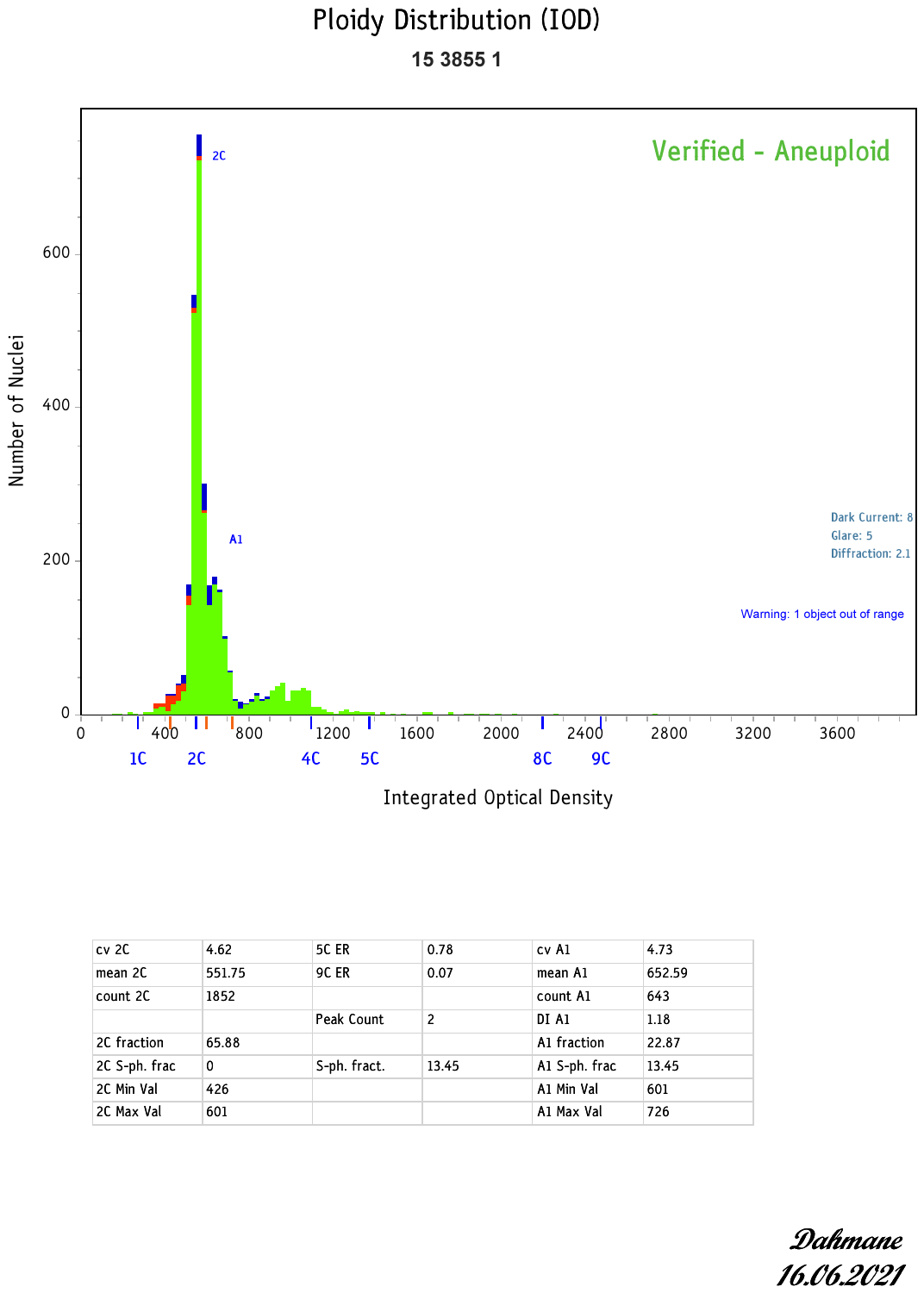

**I:** WGS_52

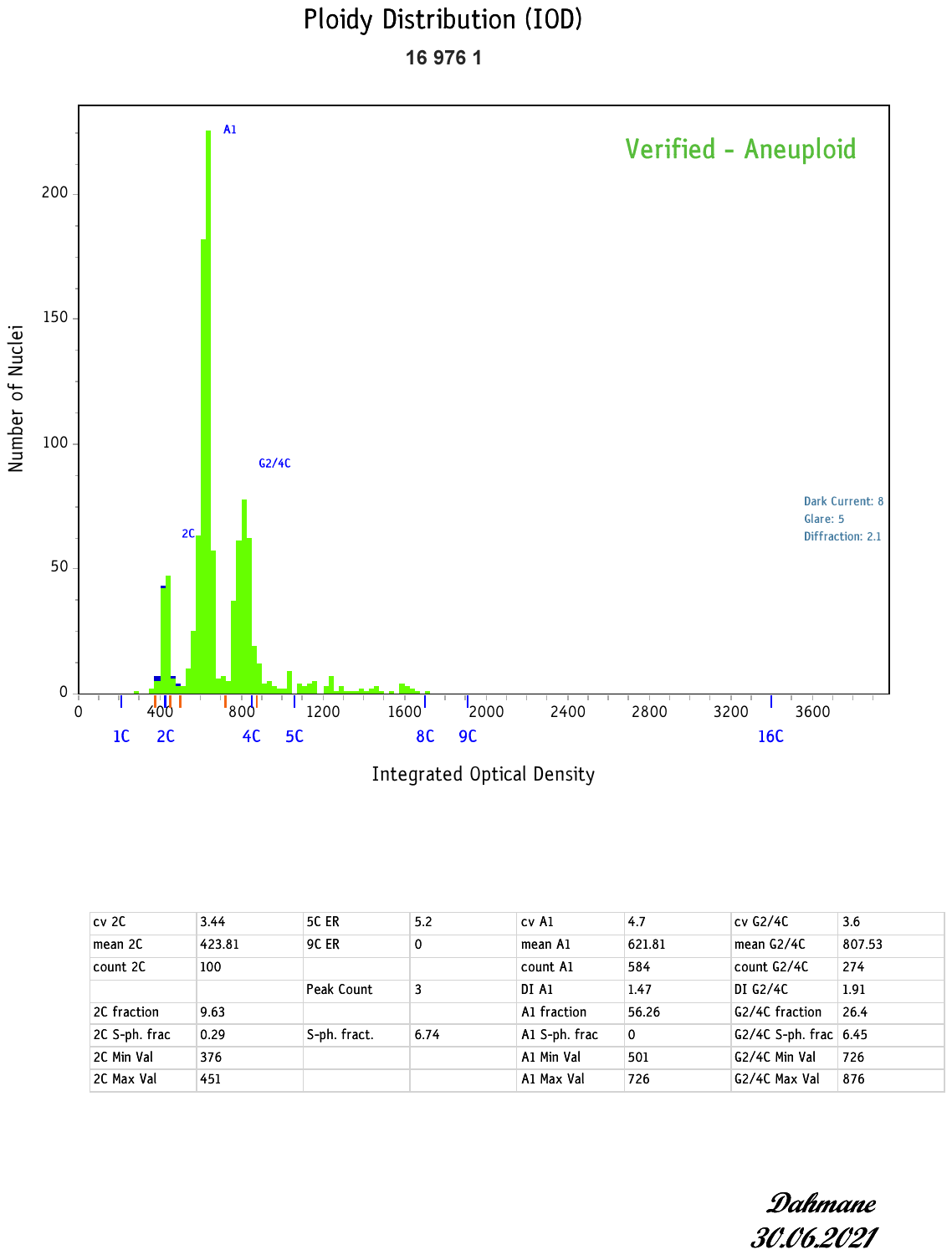

**J:** WGS_54

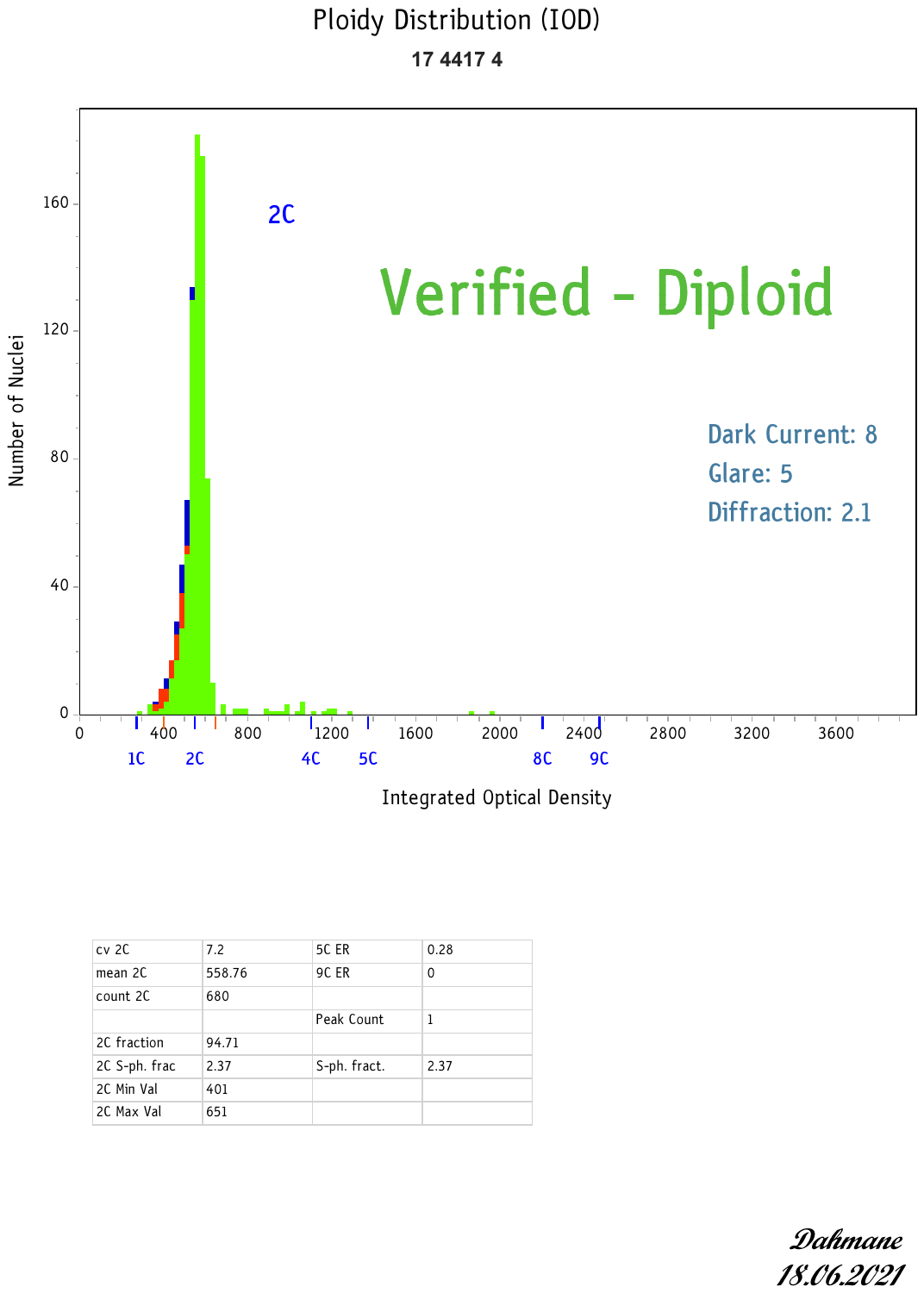

**K:** WGS_57

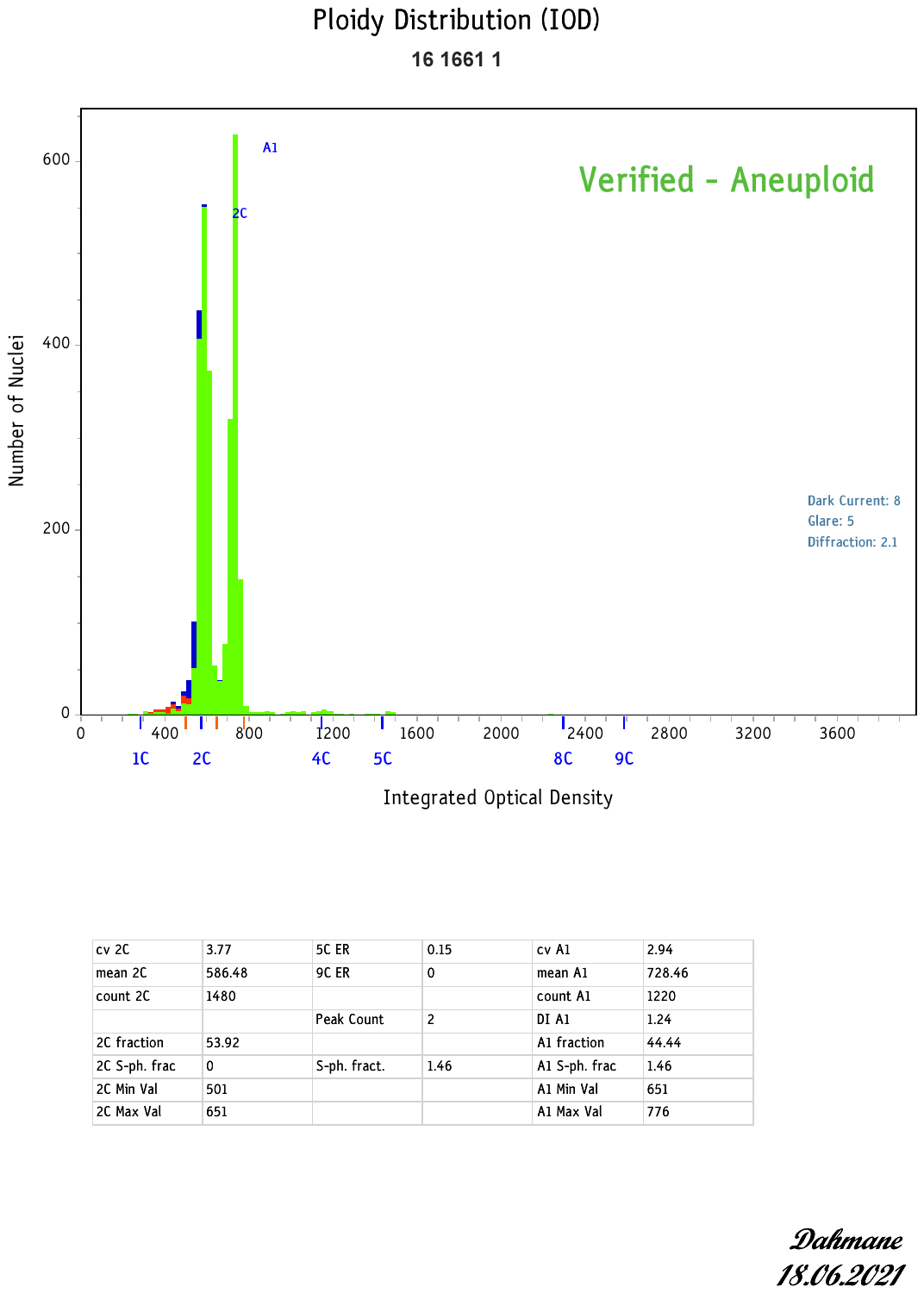

**L:** WGS_58

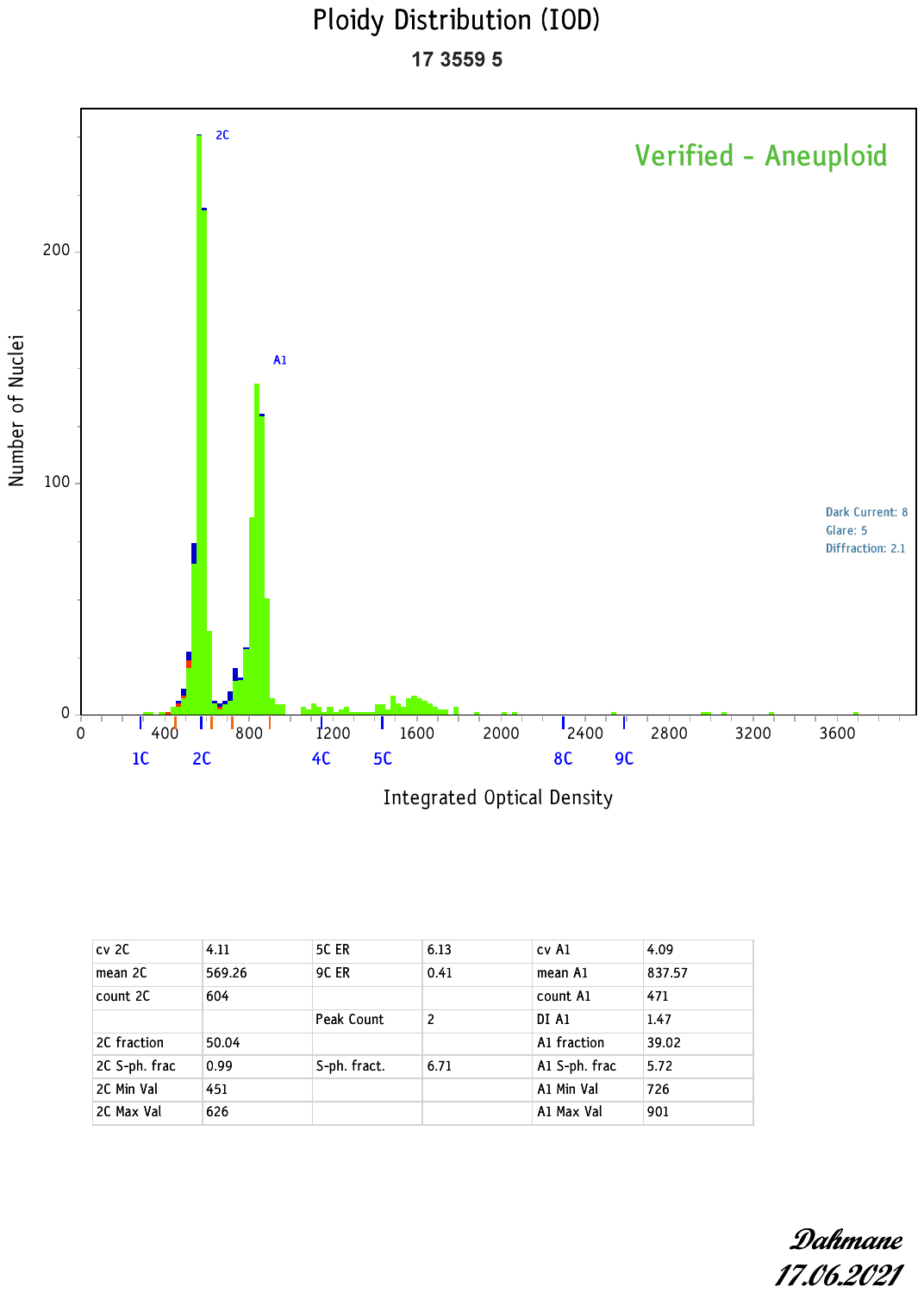

**M:** WGS_59

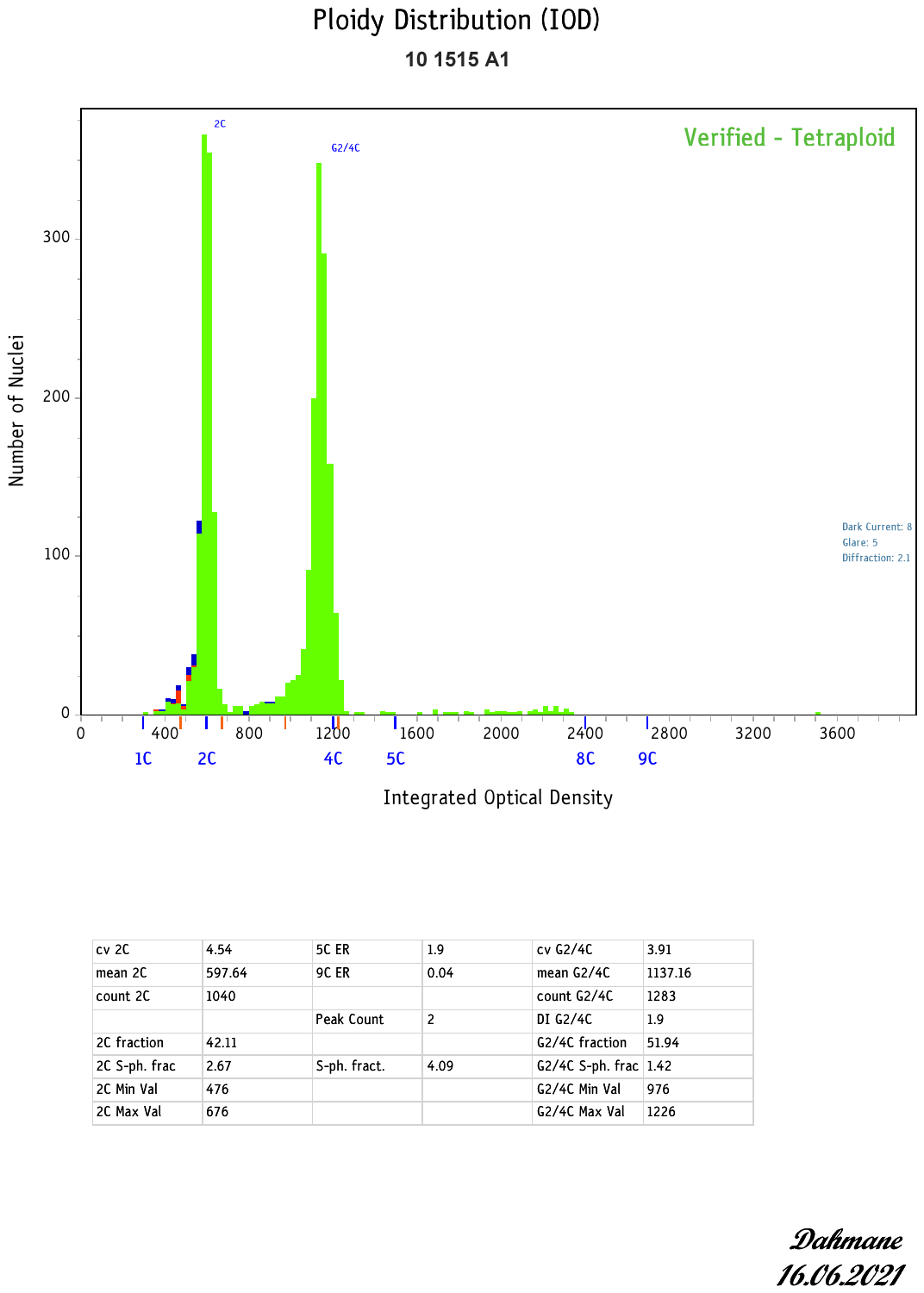

**N:** WGS_62

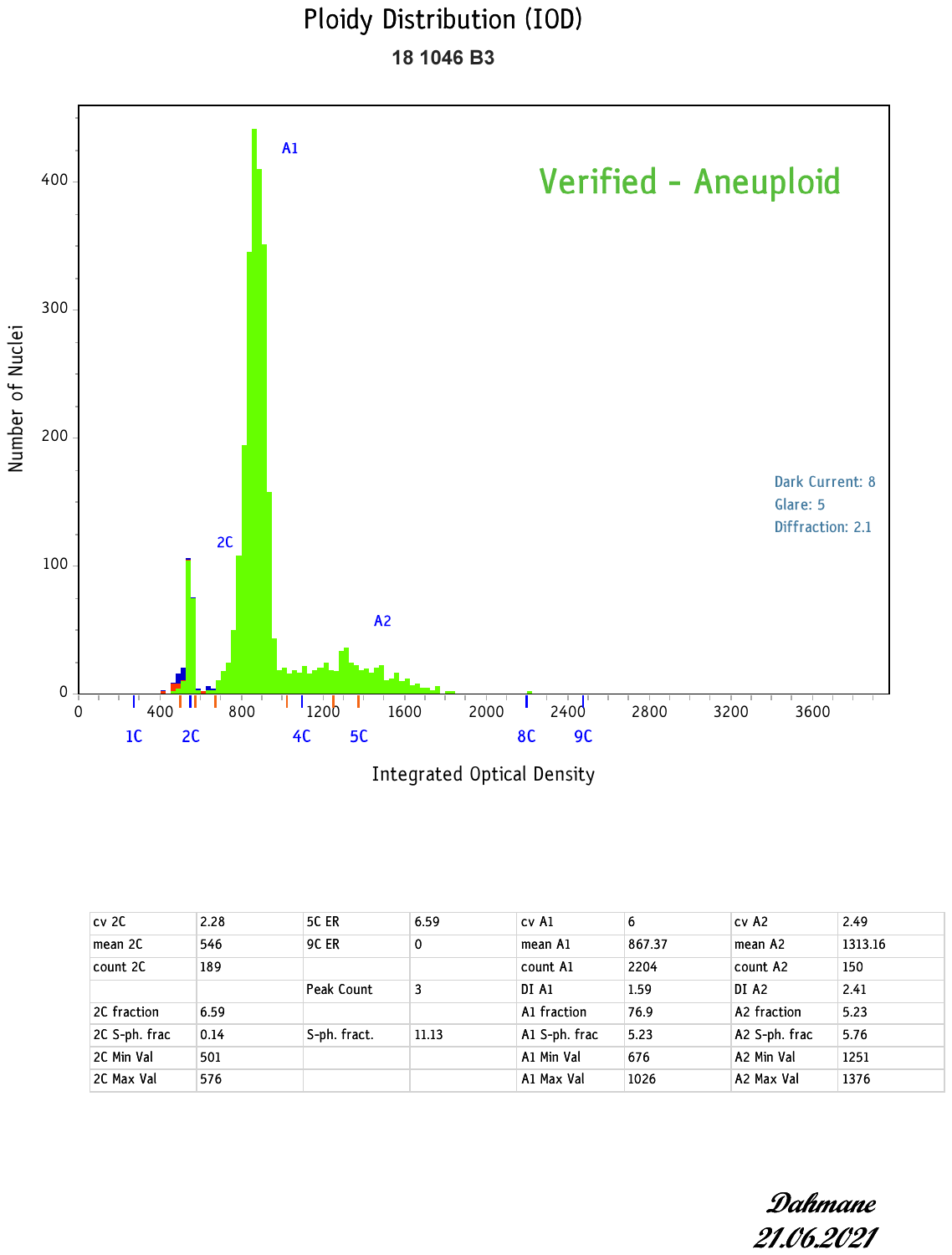

Histograms of DNA content derived from integrated optical density of each nucleus of interest (plots **A-N**). Ploidy-related parameters such as DNA index (DI) and percentages of cells exceeding 5c (5c ER) and 9c (9c ER) were also noted. Interpretation given in top right of plot.

*Note 3 – Digital PCR parameters and design tables*

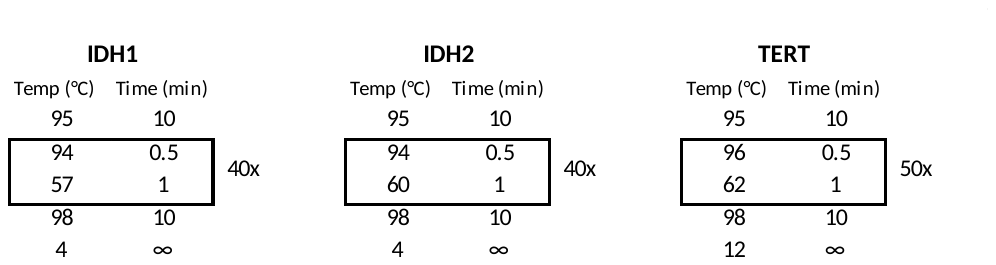

**TERT ddPCR**

| Name | Gene | For mutation in | cDNA change | Protein change | Genomic change (HG19) |
| --- | --- | --- | --- | --- | --- |
| TERT C228T _88 | TERT promoter |  | c.-124C>T | NA | 5:1253167..1295047 |
| Assay acquired from Biorad - Corless et al. 2019 | TERT C228T _88 (dHsaEXD20945488) | | |  |  |

**IDH1 ddPCR**

| **Oligos** |  |  |  |  |
| --- | --- | --- | --- | --- |
| Name | 5' modification | Sequence | 3' modification | Reaction concentration (μM) |
| IDH1-m-F | - | CCAACATGACTTACTTGA | - | 0.9 |
| IDH1-m-104-R | - | AGAGAAGCCATTATCTGC | - | 0.9 |
| IDH1-m-104SNP-R | - | AGAGAAGCCGTTATCTGC | - | 0.9 |
| IDH1-m-R132C | FAM | CCATAAGCATGACAACCTATGATGAT | BHQ1 | 0.1 |
| IDH1-m-R132G | FAM | CCATAAGCATGACCACCTATGATGAT | BHQ1 | 0.1 |
| IDH1-m-R132H | FAM | CCATAAGCATGATGACCTATGATGAT | BHQ1 | 0.1 |
| IDH1-m-R132L | FAM | CCATAAGCATGAAGACCTATGATGAT | BHQ1 | 0.1 |
| IDH1-m-R132S | FAM | CCATAAGCATGACTACCTATGATGAT | BHQ1 | 0.1 |
| IDH1-R132_WTmgb | VIC | CCCATAAGCATGACGAC | MGBNFQ | 0.1 |
| **UDG treatment?** | YES |  |  |  |

**IDH2 ddPCR**

| Name | 5' modification | Sequence | 3' modification | Reaction concentration (μM) |
| --- | --- | --- | --- | --- |
| IDH2-R172S_1F | - | CTGGCCTACCTGGTC | - | 0.9 |
| IDH2-R172S_1R | - | CCTAGTCCCTGGCTG | - | 0.9 |
| IDH2-R172S | FAM | CCATGGGCGTGACTGCCAAT | BHQ1 | 0.1 |
| IDH2-R172T | FAM | CCATGGGCGTGCGTGCCAAT | BHQ1 | 0.1 |
| IDH2-R172M | FAM | CCATGGGCGTGCATGCCAAT | BHQ1 | 0.1 |
| IDH2-R172K | FAM | CCATGGGCGTGCTTGCCAAT | BHQ1 | 0.1 |
| IDH2-R172G | FAM | CCATGGGCGTGCCCGCCAAT | BHQ1 | 0.1 |
| IDH2-R172_WT | HEX | CCATGGGCGTGCCTGCCAAT | BHQ1 | 0.1 |
| **UDG treatment?** | NO - only IDH2 R172K affected |  |  |  |

***Supplementary Tables***

*Supplementary table 1:*

Summary of data used in the analysis of the 100KGP. Columns E-H are derived from Battenberg calls. I-J are counts based on the ISAAC mutation calls from Strelka2 (see **Online Methods**). L-W are driver calls (see **Online Methods**) and columns X-Y are classifications of haploidisation, genome doubling.

*Supplementary table 2:*

Summary of clinical data and digital droplet PCR results, shown G-I for *IDH1*, *IDH2* and *TERT*. WGS driver calls shown where relevant.

*Supplementary table 3:*

Details of driver mutation SNV calls, including specific AA changes, for the 100KGP data.

*Supplementary table 4:*

Summary of methylation status for key drivers.
