## Supplementary material for "A Genetic Model for Central Chondrosarcoma Evolution Correlates with Patient Outcome": full methods

**Online methods**

*Study cohort*

DNA samples from 356 patients, with non-syndromic central conventional and dedifferentiated chondrosarcoma (CS) were included in the study following quality control as outlined below. These samples included 68 tumour-normal pairs, from patients enrolled in the Genomics England 100K Genomes Project (100KGP), on which whole genome sequencing was performed to a depth of 100x and 50x respectively (for a summary, see **Supplementary Table 1,** mutation calls **Supplementary Table 2**). Tumour sample DNA from the remaining 282 RNOH patients were subjected to ddPCR for the purpose of detecting hotspot mutations at *IDH1* R132, *IDH2* R172 and at the *TERT* promoter loci (g.1295113) as outlined below (**Supplementary Table 3**), and 64 of the WGS cases were validated using the same ddPCR assay (details below). Using the HumanMethylationEPIC and 450K beadchip, 84 were subjected to methylation arrays. Of these 6 samples were not profiled for *IDH1*, *IDH2* or *TERT*. See sample overview Venn diagram below.

*Sample overview*

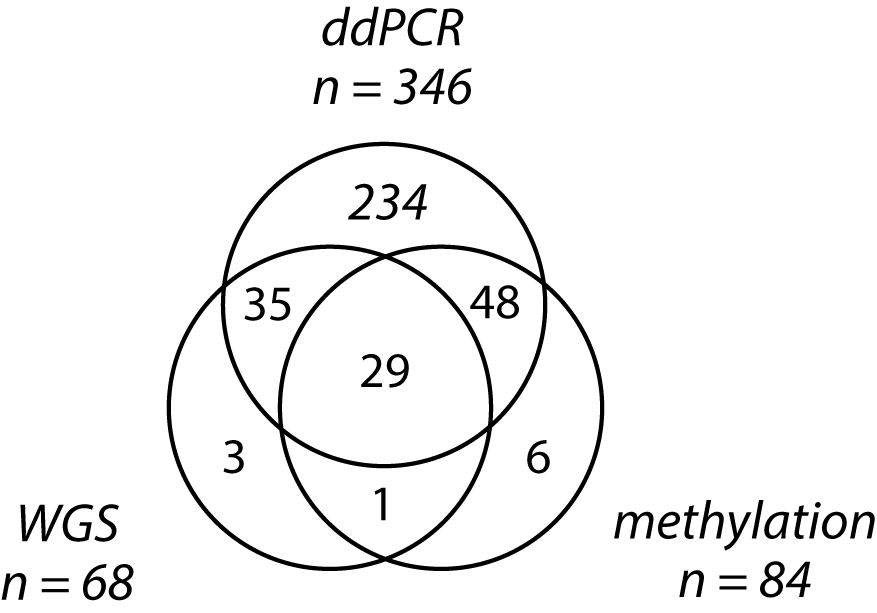

*Bioinformatic Processing within the Genomics England 100K Genomes Project*

114 CS patients were consented for inclusion in the 100KGP project. The 100KGP project involved processing all sequencing data using the Illumina-based ISAAC pipeline^1^, that leverages the Strelka2 mutation caller^2^ along with an in-house filtering step involving a Panel of Normal samples (PON) derived from the normal samples with Genomics England. An extra filter based on the GNOMAD database was also implemented. **Note:** an update to the original pipeline was performed as of version 12 (released 05/06/2021). This replaced the standard ISAAC aligner with BWA^3^, following the identification of over-clipping and subsequent issues with Structural Variant calling in the ISAAC alignments^4^. Data used in this study leveraged the BWA aligned data only.

The 123 CS genomes were subjected to quality control filtering. Only cases with confident diagnoses of central or dedifferentiated CS were taken forward. The remaining cases (n = 78) were assessed using two separate, but integrated methodologies. First, we ran Battenberg^5^ and dbClust to identify chromosome copy states, ploidy, and tumour content, and to ensure that there was a dominant clonal signal within the biopsy (in all instances there were). We then explored the single nucleotide variant (SNV) allele frequency spectrum (VAF) using the CNAqc method^6^. This reported 25 cases with low tumour content or mis-matches between the VAF spectrum and the Battenberg inferred copy states. Following up to three reruns of Battenberg using alternative parameters we were able to reset the baselines of 15 cases, resulting in a final number of 68 cases with WGS for analysis. We validated the ploidy status for a subset of these data using flow cytometry (described below). Copy gains were defined as a copy state greater than or equal to five in the context of a diploid genome, and nine or more in the context of genome doubling without prior haploidisation (genome doubling classification described below). Note that 12 of these were PCR amplified prior to WGS, hence were excluded from the mutational signature analyses, though included in the driver mutation and other analyses.

Using the Battenberg inferred copy states we inferred chromothripsis using a previously published method^7^. We also defined highly fragmented chromosome arms as those with more than 12 breakpoints present, a cutoff derived from the previous definition of chromothripsis^8^ (8-10 breakpoints across 50Kb) and examination of the distribution of breakpoint numbers.

Somatic structural variants (SVs) were identified in the 100KGP cohort using Delly v0.8.5^9^, excluding telomere and centromere regions and unplaced contigs (as per the developers' recommendations). It was not possible to obtain Delly results for three cases (WGS_ 28, WGS_34, WGS_64) due to bam format compatibility issues, leaving 65 for analysis. Variants occurring within genes were identified and annotated against the hg38 biomart gene definitions^10^ and BCFtools v1.11^11^.

As a further QC step for the WGS mutation calls, we performed the same ddPCR assay for *IDH1*, *IDH2*, and *TERT* mutations on 64 cases from the 100KGP sample set for which DNA was available. This confirmed 59 driver mutations (recall rate of 100%). We also subsequently reviewed all *IDH1*, *IDH2* and *TERT* mutations called in the 100KGP data using IGV, which were confirmed in all cases (**Supplementary Materials Note 1)**. We noted that in a single case (WGS_53) an *IDH1* mutation was reported from the ddPCR. In this situation we took the ddPCR result as a false positive, hence this case was classified as IDHwt in the final study.

*Identification of driver mutations in the 100KGP cohort and analysis using dNdS*

Driver mutations in *IDH1 R132*, *IDH2 R172*, and the *TERT* promoter were identified from their known hotspot locations^12,13^. Damaging SNV mutations were called in other genes using the default cut-offs for these tools and the amino acid substitutions annotations produced using the SIFT^14^ and POLYphen^15^ (for mutation summary see **Supplementary Table 2**). Structural variants in *COL2A1* and the *TERT* promoter region were called using Delly. *COL2A1* complex changes were manually inspected using IGV. Amplifications and copy changes, including deletions, were identified from the Battenberg calls, using the above criteria. In the case of *ATRX* and *DAXX* genes, we confirmed the WT status by manually inspecting the gene sequencing in IGV.

To examine other potential driver mutations we ran dndscv v0.0.1.0^16^ with default parameters on all reference coding sequences from Ensembl for GRCh38 using BioMart as per the instructions on the GitHub page: [http://github.com/im3sanger/dndscv/.](https://github.com/im3sanger/dndscv) We used all mutations in those coding sequences across all patient with chondrosarcoma, keeping a single PCR-Free sample per patient labelled as “PRIMARY” if available. We then looked at genes with q-value for all substitutions qallsubs_cv<0.1 (see **Supplementary Figure 2**).

*Confirmation of ploidy status by image cytometry*

We confirmed the ploidy status in 14 of cases using image cytometry^17^. One 50 *µ*m section of FFPE tissue was cut from a tumour-rich block that was selected by a pathologist. Nuclear monolayers were prepared by dewaxing with xylene and ethanol, then digesting the curls using proteinase XXIV (Sigma-Aldrich, UK) for 2 hours at 37°C. The lysate was strained through a 40 *µ*m nylon mesh cell strainer (BD Biosciences, USA), producing a nuclear suspension. This was spun onto charged Superfrost Plus microscope slides (electrostatically permanently positive charge, VWR, UK) at 225 g for 5 min. The slides were dried for 1 hour and then placed in 5M HCl for 1 hour. The slides were then stained with Feulgen–Schiff reagent using standardised methodology. The stained monolayers were then analysed using The Fairfield DNA Ploidy system (Fairfield Imaging, UK). Optical density and nuclear area were measured, and the integrated optical density of each nucleus was calculated after correcting for the background. Segmentation software selected at least 1000 whole nuclei and sorted into four separate cell galleries: nuclei of interest for measurement included lymphocytes, plasma cells and fibroblasts. The lymphocytes were used as reference cells to determine the position of the diploid peak (2c). The galleries were then edited manually to discard any overlapping nuclei. The integrated optical density of each nucleus of interest was calculated and a histogram of DNA content produced. Ploidy-related parameters such as DNA index (DI) and percentages of cells exceeding 5c (5c ER) and 9c (9c ER) were also noted. Histograms were analysed according to European Society for Analytical Cellular Pathology guidelines^18^ (for results see **Supplementary Materials Note 2).**

*Identification of genome doubling and partial haploidisation*

Using the equation defined from the PCAWG study ( 2.9 – (2 * %homozygous) <= ploidy, see <https://github.com/gerstung-lab/PCAWG-11>) and our Battenberg calls, we classified samples as genome doubled or non-genome doubled. As means of validation we created a classification method to call both genome doubling (GD) and haploidisation (HP) based on the the Mclust R package^19^. We used percentage of the genome at copy neutral LOH state (cnLOH), and percentage of the genome aberrant (PGA) as input. Both methods returned the same 23 cases classified as GD (**Supplementary Figure 4**). Reviewing scatter plots of the ploidy, percentage LOH, and percentage genome aberrant (PGA) for these, data revealed a clear demarcation between cases that had more than 50% of their genome at cnLOH state. The characteristic widespread cnLOH can be explained briefly by a partial haploidisation event preceding GD, as this yields a diploid to triploid genome, with high levels of cnLOH (>50%). HP cases were therefore defined by a 50% cut-off in both copy neutral LOH and PGA. The average ploidy of GD, non-HP cases was significantly different to the non-GD classified by this method (ploidy non-GD = 2, GD (non-HP) = 3.2, p = 3e-9).

*Digital Droplet PCR*

We performed ddPCR on 401 cases of CS following previously a published method^20^. Briefly, tumour DNA was analysed on the QX200 Droplet Digital PCR System (Bio-Rad, CA, USA). 20 μL reactions consisted of up to 9 μL DNA, 10 μL 0.02x Supermix for Probes (no dUTP; Cat.No 186-3023, Bio-Rad, USA), 18 mM forward and reverse primers, 0.05 mM probe, and nuclease-free water. Droplets were generated on the QX100 Droplet Generator (Bio-Rad), which then underwent either 40 or 50 cycles of PCR (T100 Thermocycler, BioRad; 95⁰C for 10 min, 40 cycles of 94⁰C for 30 sec/50 cycles of 96⁰C for 30 sec, XX⁰C for 1 min (XX = assay specific, **Supplementary Materials Note 3**) prior to reading on a QX200 droplet reader (Bio-Rad). Droplets were read with either the FAM/VIC or FAM/HEX channels setting provided by the QuantaSoft 1.7.4 software package (Bio-Rad). Droplets were inspected visually and called as ‘mutant only’, ‘WT only’, ‘double-positive’ or ‘template negative’. A positive control, WT DNA and no-template control were included in each run. The positive control was a patient’s sample with a mutation verified via whole genome analysis, and the WT sample was a commercially available pooled sample of human placental DNA (BioLine, London, UK). The inclusion of the WT and no template control (H2O) in each assay run was used as a measure of background error rate. The plots were visualised in all cases. Each DNA sample was run in duplicate (up to 300ng tumour DNA loaded per well) on each assay run. Tumour samples were deemed to be mutant-positive if they met a minimum threshold of 3% “mutant only” droplets with over 10,000 droplets also called. Primer designs and other information are given in **Supplementary Note 3.**

*Timing genome doubling events*

It is possible to time chromosome changes using splits in the variant allele frequency, as previously reported^21^. We implemented a genome doubling timing method as previously described^22^. Briefly, a binomial mixture model was used to estimate the proportions of clonal SNVs on one and two copies in regions of the genome with a clonal major copy number of two. The distributions on the proportion of mutations on two copies were converted into clonal mutational time separately for genomic regions with 2+0/2+2 and 2+1 copy number states. These two distributions were then combined to form an overall distribution for the timing of the genome doubling.

*Analysis of mutational signatures*

96 channel single-base substitution *de novo* mutational signatures, subsequently matched to COSMIC reference mutational signatures^23^, were extracted from Strelka-called SNVs using SigProfilerExtractor version 1.1.3^24^ with default parameters.

*Methylation data protocol and analysis*

500 ng of DNA from frozen tumour samples were bisulfite converted using Zymo EZ DNA methylation Gold kit (Zymo Research Corp.Irvine, CA, USA) and hybridised to the Infinium HumanMethylationEPIC beadchip arrays (Illumina, San Diego, CA). The generated methylation data were analysed using the ChAMP R^25^ normalised using BMIQ and hierarchical clustering plots were constructed using the ‘pheatmap’ R package^26^ (see **Supplementary Figure 3**).

*TERT* promoter methylation status was determined by looking at the methylation status of the cg11625005 probe as reported previously^27^. Raw DNA methylation data files have been deposited in the ArrayExpress database at EMBL-EBI ([www.ebi.ac.uk/arrayexpress](http://www.ebi.ac.uk/arrayexpress)) under accession number E-MTAB-11031.

*A note on statistics and mathematical analysis*

In all group comparison situations, such as *IDH1*, *IDH2*, IDH-WT cases, with or without *TERT* mutations, we used Fisher test statistics as implemented in R (testing both 3X2 and 2x2 contingency tables). Distributions of data, as seen in the tests of timing for genome doubling (Figure 1E) was performed using Wilcoxon tests, again implemented in R. Linear regressions (Figure 3A) were also implemented using the standard R methods. The cox proportional hazard model and Kaplan-Meyer analyses were performing using the *survminer* package^28^. For the chromosome arm frequency comparisons, we used fisher tests and the Bonferroni multiple testing correction.

*Power calculations and sampling analysis*

High grade *IDH2*-mutant tumours with outcome data are limited in numbers compared to the other groups (n = 38) and likewise *TERT*-mutant cases are present at low frequency in the *IDH1* group (n = 116, *TERT* mutant, n = 36). For these reasons we performed a power calculation to increase confidence in our reported outcome statistics. We used the downloadable version of G*Power^29,30^ for this purpose, and ran the "Post hoc power calculation" module for *IDH1* and *IDH2* data. For a sample size of n = 38, one degree of freedom (*TERT* wt or *TERT* mutant), an alpha value of 0.05, and a frequency of *TERT*-mutant of 68%, as per the *IDH2* data, a power of 0.86 was calculated for an effect size of 0.5, and 0.69 for an effect size of 0.4. The actual effect size in the *IDH2* outcome data was calculated as 0.04 between *TERT* mutant. and *TERT* wild type. For the same calculation in the *IDH1* group, using a sample size of n = 116, and a frequency of *TERT* mutant of 31%, a power of 0.99 was calculated for an effect size of 0.4. The effect size for these data was 0.38, and a power of 0.97 was calculated for these data. We therefore considered these data sufficient to draw our conclusions.
